# Robust Feature Selection for Cancer Microarray Data Using a Hybrid mRMR and Binary Lion Optimization Algorithm

**DOI:** 10.1101/2025.10.07.25337478

**Authors:** Bibhuprasad Sahu, Amrutanshu Panigrahi, Abhilash Pati, B K Madhavi, Janmejaya Mishra, Ram Kaji Budhathoki, Saurav Mallik

## Abstract

Microarray cancer datasets are characterized by a large number of irrelevant, redundant, and noisy features, which can severely hinder the accuracy and efficiency of classification algorithms. Feature selection, as a crucial branch of feature engineering, aims to enhance classification performance by identifying and retaining only the most informative features. However, feature selection is an NP-hard problem, where conventional search strategies are often prone to premature convergence and local optima, resulting in increased computational burden. To address these challenges, global metaheuristic algorithms have been widely explored. The recently proposed Lion Optimization (LO) algorithm has shown promising results for continuous optimization problems, yet its design is not inherently suited for discrete feature selection tasks. To overcome this limitation, a binary variant of the LO algorithm, termed Binary Lion Optimization (BLO), is introduced for wrapper-based feature selection in microarray cancer data analysis. In this work, the Minimum Redundancy Maximum Relevance (mRMR) criterion is first employed as a filter method to identify an initial subset of relevant features, thereby reducing search complexity. The refined feature subset is then optimized using the BLO algorithm to achieve improved classification outcomes. The proposed mRMR-BLO framework was evaluated on several widely recognized cancer microarray datasets and benchmarked against four state-of-the-art binary optimization algorithms. Experimental results demonstrate that mRMR-BLO consistently identifies smaller yet highly discriminative feature subsets, while achieving competitive or superior prediction accuracy. These findings highlight the potential of mRMR-BLO as an effective and robust tool for high-dimensional microarray cancer classification.

## 1. Introduction

There are several areas, such as social media, financial services, telecommunications, etc., that are continuously churning out and collecting data due to the rapid improvement of Internet technology. For data extraction to be more purposeful, data collection and cleansing are necessary due to the significant amount of relevant and irrelevant data existing in these domains. Feature selection, which is recognizable and widely used for data cleansing, is considered to be one of the most efficient techniques and is applied in multiple fields, including data mining, medicine, and even pattern recognition. This step of feature selection includes duplicate feature elimination and constructing an optimal subset of features that will be the most fitting in that context. This process includes a search strategy and evaluation criteria: the first strategy is designed to measure the quality of feature subsets to guide the search, and the second strategy explores the search space to find the best feature subset. At this stage, three methods are applied to evaluate the criteria to determine the selection of the feature subset: filter-based, wrapper-based, and embedded-based procedures. Filter-based is one feature selection method that uses statistical information as a basis for assessing feature subsets. The wrapper-based method utilizes a classifier to assess feature subsets. When the classifier is in the training stage, he is also allowed to determine the subset of features; this is done using the embedded-based method [1].

As expected, although the wrapper-based approach works well, it demands more computing resources than the other two techniques. Selection of features is a very difficult optimization problem, and it is combinatorial at the same time. The goal of this process is to find the best feature subset with the help of different search techniques, such as exhaustive, random, or heuristic search. The exhaustive search approach allows optimal selection of feature subsets of 2^*N*^ subsets that can be created from N features [2]. Nevertheless, this approach encounters severe computational costs as the number of features increases. An alternative is a random search where new candidate subsets are iteratively produced until a desirable subset is achieved. The third search method is a heuristic that broadly covers both sequential forward and backward selection methods. These have recently been implemented in a more generalized manner, which makes them easier to use; the variability of which is highly desirable. The principles of these algorithms allow them to be classified into four categories: (1) evolution-based algorithms, (2) physics-based algorithms, (3) swarm intelligence-based algorithms, and (4) human-based algorithms. This category comprises algorithms that draw from Darwinian processes of natural selection. A genetic algorithm (GA) is a well-known evolution-based algorithmic technique that stages biological evolution processes [3]. Like genetic algorithms (GA), the differential evolution (DE) algorithm is composed of three elements: selection, crossover, and mutation. The emergence of new members in DE is determined through the cooperation and competition between the individuals. Novel or improved methods for creating high-grade techniques or solutions to the problem are borne out of the need to counter the no free lunch theorem of optimization problems. The arbitrary population is then subjected to the objective function of performing a fitness test in which the values of all the features are computed, and the accepted values are selected. This step is done with the K-nearest neighbours (KNN) classifier. Based on the value of the fitness function, a change is defined. This operational plan is repeated until the criteria for stopping the algorithm are met. These processes in a cycle give the optimum determination of features that can be processed by the machine learning classifiers. The capacity of the algorithm plays a significant role in making such a fundamental mark on the single-objective binary optimization problem. It was better than other highly sophisticated optimization algorithms that are based on the same systems. Due to the effectiveness of the algorithm in solving the intricate challenges of continuous optimization systems, the question we set out to answer was whether it was possible to achieve high-quality solutions using these operators and optimization strategies for binary optimization problems cite seyyedabbasi2025v.

This work aims to determine the effectiveness, benefits, and consequences of creating and applying a binary LO technique via meticulous testing on various complex datasets. In the beginning, this approach was never utilized to solve the feature selection problem; however, the productivity of the approach prompted this binarization. So as to address the bounded-continuity facet of the feature selection problem, we propose a binary form of the LO. The KNN classification is used to evaluate the effectiveness of the chosen set of characteristics. In my opinion, the following are the most important contributions of the work, as stated:

- A novel binary feature selection algorithm based on the Lion algorithm was proposed. The performance of the BLO feature selection algorithm has been assessed using a range of computational analysis metrics.
- Eleven cancer datasets with varying sizes and dimensions were evaluated for the effectiveness of the proposed feature selection algorithm.
- mRMR-Binary LO is compared with several widely used feature selection techniques, and their effectiveness is measured using three different classifiers.
- The proposed method is compared with various existing methods to test its efficacy.

Machine learning models tend to have to deal with the decrease in accuracy in highdimensional data, which is why feature selection is crucial to not only improving predictive accuracy but also ease of computation. Filtering procedures pick out the features confidently and efficiently that are very relevant to the target variable and are resistant to easy redundancy. However, these methods are not influenced by any classifier and will not be able to identify higher-order interactions between variables, which can be used to strengthen predictive ability. This advantage may be offset by the fact that in a wrapper-style candidate feature subset evaluation, candidate features are evaluated as a component of a specific classifier and thereby may include intrinsic feature dependencies and may also result in better predictive capability. However, BLO, in some cases, is computationally expensive and is sometimes prone to unstable optimization in large search spaces. In continuation of these observations, it is possible to introduce a hybrid model consisting of mRMR and BLO that is assembled in sequential chronological order. First, mRMR brings about a reduction of the feature set by removing duplicate or irrelevant features, therefore, reducing the search space. The final pruned set is then explored on BLO, and thus, a more focused model-specific search is done to find the best conjunction of features. The benefit of this sort of synergistic integration is that it has all the benefits of statistical relevance-filtering of mRMR and the optimization power of BLO, and helps in compensating for the drawbacks of both. The introduced methodology is a systematic and computationally feasible way of feature selection, a setting, as far as we are aware, previously studied with respect to BLO. The empirical analysis shows that all other benchmark models are outperformed by the proposed hybrid one with respect to classification accuracy. Therefore, it can be assumed that the current work makes a methodological contribution of substance and offers practical benefits when it comes to practice in high-dimensional feature selection.

The rest of the study is described here. Section 2 presents a survey of the literature, followed by materials and methods in Section 3. Section 4 presents the results of the experiments and the performance comparison. Conclusion and future scope of the study are described in Section 5.

## 2. Literature survey

In cancer datasets, the selection of features is crucial to building accurate classification models. Binary metaheuristic algorithms may be useful in improving feature selection in high-dimensional data. This review of the literature presents the latest information on cancer feature selection, binary metaheuristic algorithms, and their classification accuracy results. Machine learning algorithms have been shown to improve early cancer detection. In classifying breast cancer using risk factors, Alfian et al. [1] used a support vector machine SVM and an exceptionally random tree classifier. The authors noted that their model attained 80.23% accuracy, which is better than the other machine learning models. Rahman and Muniyandi [2] investigated feature selection techniques to increase the accuracy of colon cancer classification. Their feature selection used an artificial neural network (ANN) that utilizes a best-first search and achieved 98.4 percent accuracy. The study highlighted the importance of the selection of relevant characteristics to improve the classification performance, especially in cases where the number of characteristics is considerably larger than the number of samples. Banati et al. [3] proposed the binary form of the peacock feature selection algorithm, which is a binary metaheuristic method, to validate feature selection. With the 34 benchmark datasets, bPA achieved the highest accuracy in 30 classification datasets and the selection of feature subsets in 32 datasets in which feature dimensionality was reduced by 99.80%. The findings indicated that bPA is capable of improving feature selection methods used in cancer dataset classification problems. Seyyedabbasi et al. [4] analyzed large biomedical datasets focusing on the important aspect of feature selection. The bAPO algorithm improved the search and exploitation of the solution space through adaptive population dynamics. The bAPO algorithm achieved better fitness values and classification accuracy than the KNN and SVM classifiers in 14 significant biological datasets. Feature selection helped in model building, which proves the effectiveness of metaheuristic methods in this area. Gafar et al. [5] applied the RBAVO-DE(Relief Binary African Vultures Optimization based on Differential Evolution) method for RNA-Seq gene selection and found, with the help of differential evolution, that they can overcome high dimensionality problems. To their surprise, they achieved 100% classification results along with an astonishing 98% reduction in feature size on 22 cancer datasets. The results provided evidence that RBAVO-DE is undoubtedly helpful in more efficient gene selection, which aids cancer researchers in finding the genetic markers.

Ahmad Zamri et al. [6] used and evaluated a novel microarray feature selection method based on a Kalman filter with mutation (SKF-MUT). They picked the most useful informative features in high-dimensional datasets to increase the classification performance of the ANN model. Skf-mut achieved classification accuracies between 95% and 100%, demonstrating the effect of feature selection on cancer diagnostics machine-learning algorithms. Halder et al. [7] proposed GFS-Net, a grey wolf optimization (GWO) based oral cancer classification model with deep feature selection. This model utilized a convolutional neural network(CNN) for feature extraction and GWO for feature selection. GFS-Net obtained classification accuracies of 92.86% and 93.94% on two oral cancer datasets. Akinola et al. [8] have shown that metaheuristic algorithms for evolutionary, swarm intelligence, physics-based, and humanintelligence-based multiclass feature selection are very efficient. These methods construct appropriate feature subsets that enhance the performance of the classifiers. Hameed et al. [9] showed that cancer datasets can be used to test the performances of binary metaheuristic algorithms such as Binary particle swarm optimization(BPSO), GA, and Cuckoo Search(CS). Their three-phase hybrid approach, which uses a PPMCC pre-processing filter, increases speed and accuracy at the same time. BPSO achieves higher classification accuracy as compared to GA and CS, even though CS is able to achieve lower computing costs, and fewer genes are selected. The use of appropriate metaheuristic algorithms in filter-wrapper models for the selection of cancer genes greatly influences feature selection research. One of them is binary variations of optimization algorithms, like Hunger Games search optimization. The process of feature selection within classification tasks gets better with the utilization of *V* and *S* shaped transfer functions as they transform continuous optima into binary ones. According to studies by Manjula Devi et al. [10], BHGSO-V outperformed with regard to 16 UCI datasets, achieving an average classification accuracy of 95% while at the same time recording minimal computing times. These results further affirm that the application of Hybrid Binary Metaheuristic Algorithms can improve the outcomes of Feature Selection. To tackle the issue of discrete search spaces in FS problems, the African Vulture Optimization Algorithm has been modified into its binary version, BAVOA. The BAVOA S2-BAVOA, working with eight transfer functions, surpassed other traditional binary metaheuristics in feature selection, F1 measure, and classification accuracy on fourteen benchmark datasets. According to Balakrishnan et al. [11], S2-BAVOA was able to outperform 15 other Traditional Binary Metaheuristic Algorithms using 8 transfer functions. These results confirm that hybrid techniques are useful for feature selection problems. This approach is actually very useful for these cases. Khan and Nisha [12] combine metaheuristic methods for FS and achieve greater classification accuracy and speed of convergence in the hybrid approach. GWO excels in many functions, but local optima tend to be a pitfall for it. Al-Wajih et al. [13] proposed HBGWOHHO, which incorporates a hybrid mixture of GWO and Harris Hawks Optimization(HHO) to achieve a better balance between exploration and exploitation. The method fulfils feature selection criteria by using a sigmoid transfer function, which transforms continuous search spaces into binary ones. The suggested method shows superiority over BGWO and BPSO on 18 UCI benchmark datasets in terms of accuracy, feature size, and time taken to compute. For classification feature selection, Bezdan et al. [14] proposed a hybrid metaheuristic approach that integrates the Brain Storm Optimization (BSO) algorithm with the Firefly method. This wrapper-based approach was applied to 21 datasets, including the coronavirus disease dataset. This hybrid approach reduced the feature subset and outperformed eleven other metaheuristic classification algorithms, demonstrating effectiveness in real-world problems. Social media disinformation reasons the need for Text classification techniques. Zaheer et al. [15] created a meta–heuristic filter wrapper feature selection based on Binary Dragonfly for categorization of false news. The model performed exceptionally well and showed robustness across multiple datasets for solving categorization problems utilizing binary meta-heuristics. The use of the Internet of Things IoT provides new challenges in security, especially for intrusion detection systems. Geetha et al. [16] devised a classification model based on feature selection that utilizes a Chaotic Vortex Search (CVS) for feature selection and a Fast-Learning Network (FLN) for classification. Their studies conducted on the CIC IDS-2017 and BoT-IoT databases were astonishingly accurate and specific, further reinforcing the role of hybrid models in the security of IoT. This paper demonstrates how binary metaheuristics can be applied in trustworthy severe domains. In another research for polarimetric synthetic aperture radar (POLSAR) image categorization, Sadeghi et al. [17] used hybrid MOBChOA and deep convolutional neural networks - only this approach of classifying features yielded the highest accuracy. This particular method argues for the applicability of hybrid approaches toward challenging picture classification tasks and emphasizes the role of binary metaheuristic algorithms for deep learning feature selection.

Moosavi et al. [18] have expanded the discussion on feature selection with the Hybrid Sine Cosine Firehawk Algorithm. This method introduces a new metaheuristic approach in which the minimization of data set variance is incorporated into the cost function as a method to overcome K-Nearest Neighbor (KNN) limitations. It is possible to preserve data quality with greater efficiency by combining hybrid metaheuristics, as demonstrated by enhanced feature selection performance on 22 UCI datasets. In [19], the authors proposed an enhancement to the shuffled frog leap algorithm by including a chaos memory weight factor, a complete balance group strategy, and an adaptive transfer factor to search for subsets in the features of high-dimensional biomedical data that optimize significance and accuracy while ignoring noisy and irrelevant features. The hybrid design implements the filter technique as a conditional mutual information maximization within the wrapper technique, which operates as a genetic algorithm to improve performance metrics in classification tasks and accelerate the search for key features. This method will be referred to as FWFSS. To eliminate irrelevant features and identify the relevant biomarkers, the wrapper method, as a genetic algorithm, applies the Naïve Bayes (NB) classifier as the fitness evaluation tool [20]. The authors introduced a hybrid filter-wrapper gene selection method with minimum redundant maximum relevance (MRMR) as the filter step and the flower pollination Algorithm (FPA) as the wrapper step. MRMR was used to extract the most critical genes from gene expression data, and FPA works to identify the optimal informative gene subset from the reduced set generated by MRMR [21]. In this research, gene selection is addressed using a two-tier approach: the filter stage that applies the MRMR technique and the wrapper stage where BA and SVM are utilized [22]. To address problems related to excessive dimensionality and overfitting, the authors in this study created a novel gene selection technique using the mRMR method combined with teaching-learning-based optimization (TLBO) for precise cancer prediction. MRGMR is first implemented in the proposed method to identify the most discriminative genes within the original feature sets. After this step, TLBO with Opposition-Based Learning is applied to the reduced feature set to further refine the feature set that can aid in classifying the cancers. In addition, activation functions for more efficient gene selection are introduced, which serve the purpose of transforming the search space from continuous domains to binary domains. In the proposed method, the Support Vector Machine (SVM) is utilized as a fitness function to estimate the predictive accuracy and classify cancer, enable the selection of relevant features, and improve the accuracy of cancer diagnosis [23].

In [24], the model has been reported to utilize the feature selection algorithm Minimum Redundancy Maximum Relevance (MRMR) as its basis. The Whale Optimization Algorithm (WOA) is implemented on the featured dataset to optimize the number of features selected without losing relevance to the microarray data. To overcome the specific difficulty encountered with the analysis of biomedical data, an innovative feature selection approach based on an enhanced binary clonal flower pollination algorithm is developed to remove irrelevant features while guaranteeing the high accuracy of disease classification. Incorporating the absolute balance group strategy and adaptive Gaussian mutation helps increase the diversity of the population and improve the search performance. The classification accuracy is assessed using the KNN classifier [25]. A new enhancement hybrid feature selection method has been created using the Modified Binary Aquila Optimizer with Time-Varying Mirrored S-Shaped Transfer Function (MBAO-TVMS) that is capable of obtaining suboptimal partitioning of relevant genes that can optimize the gene feature selection problem. Adding mRMR as a filtering method, the hybrid method aims to eliminate redundancy while maximizing relevance, whereby it ranks the genes first and later executes the two steps – identifying top-ranked discriminative genes and selecting the most discriminative ones - in a wrapper approach in the second phase. The continuous version of the AO is transformed to its binary form using TVMS, and a mutation mechanism is added into the binary AO to enhance the ability of the algorithm to evade local optima and strengthen its global search ability [26].An effective tumor identification system is implemented based on a hybrid strategy using multiple filters combined with a recursive flower pollination algorithm (MFI-RFPA) for gene selection, aiming at optimal classification performance with minimal gene selection. In MFI-RFPA, multiple filters aligned with different measures are aggregated, reinforcing the filtering process. In the second step, an optimal subset of the leftover genes is searched by MFI-RFPA using a recursive flower pollination algorithm (RFPA) [27]. To tackle cancer classification problems and improve their performance, the authors proposed a novel feature selection method that combines Random Drift Optimization (RDO) with XGBoost to address the issues of dimensionality reduction challenges [28]. Authors in [29] used Darwinian theories with the binary GWO algorithm to optimally select the subset of features in the dataset of cancer. The tangent hyperbola method, based on information gain (IG) and the MBKH algorithm, proposed a new hybrid approach to achieve optimal feature subset selection. As a result, information gain as the statistic permits to provide a useful candidate subset for the wrapper step and accelerates the convergence rate of the wrapper section, assuming the MBKH structure. In this work, the adaptive transfer factor and chaos memory weight factor are used for fine-grained searching [30]. Also, in reference [31], a new extension of the Multi-Objective Fish Optimization Algorithm (MFOA) called modified Moth Flame Algorithm (mMFA), with the mMFA combined with the maximization of mutual information (MIM), to tackle the gene selection problem in microarray data classification. Authors in [32], developed a novel model called BIMSSA. In this study, the pipeline concept is implemented with the Salp Swarm Optimization(SSA) algorithm to select the efficient features from cancer datasets. A hybrid model with a combination of Cuckoo and spider monkey optimization (SMO) algorithm is proposed, followed by mRMR to select the high-ranked features from the original datasets [33]. A hybrid model called CRO-SA is proposed to handle the high-dimensional datasets. In this study, SA is used as a local search strategy to maximize the efficacy of Coral Reefs Optimization (CRO) [34]. Similarly, in [35], the Symmetrical Uncertainty (SU) algorithm is a local search with the Reference Set Harmony Search Algorithm to maximize the searching mechanism. Other authors in [36] and [37] proposed novel filter-wrapper models for significant feature identification. In [38], the authors proposed an advanced version of the lion optimizer to enhance the local search for easy identification of biomarkers from the dataset using SVM. Authors in [39] proposed a hybrid model by integrating lion optimizer with firefly algorithm for feature selection. A deep learning-enabled lion optimizer is proposed to handle the Network Intrusion Detection System by collaborating with an attention-based bi-directional long short-term memory approach [40]. Similarly, authors in [41] presented a parallel Lion Optimization model to select biomarkers from a microarray cancer dataset. Authors in [42] proposed a deep learning enabled hybrid model for the grading of mango fruit. The lion algorithm is preferred by researchers for optimal feature selection. It motivates us to propose an mRMR-enhanced binary lion optimization model for cancer prediction.

## 3. Materials and Methods

### 3.1. Lion Optimization Algorithm

This Algorithm is designed according to the hunting and living nature of lions. Basically, lions lived two types of life, such as Pride and Nomadic. In Pride, the lions live with five females and their cubs. But in Nomad, the lions separated from the pride and started looking for a partner. During food search, lions, in cooperation with family members, attack the prey from a different direction. The coordination in hunting is a great skill that the lions have executed to get their food. During mating, male lions prefer multiple lionesses when they are in hit. The territory of the lions is defined by their urine smell. The LO algorithm is a population-based metaheuristic algorithm designed on the hunting, living, and mating nature of lions [43]. The schematic presentation of LO is presented in Figure 1.

**Figure 1:**
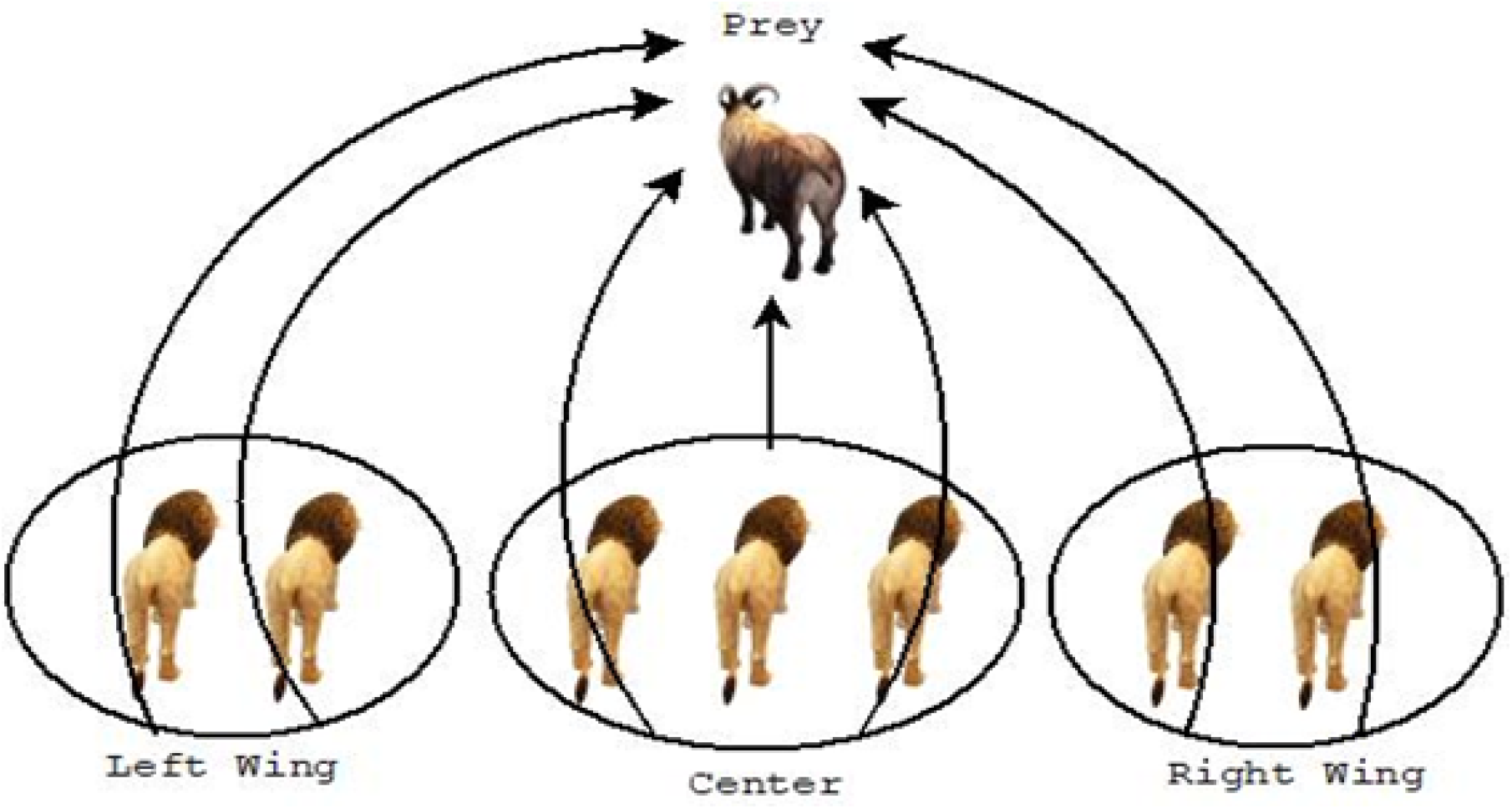
Schematic presentation of Lions in the population.

#### 3.1.1. Initialization of Population

The population (L) of the lion search is randomly generated, where each solution in the search space is known as a lion. The dimension of the population search space is presented as (N). Equation 1 presents the mathematical presentation of the population.

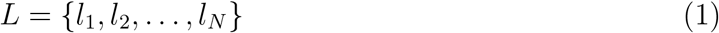

The fitness value of each Lion is calculated using the cost function presented using Equation 2.

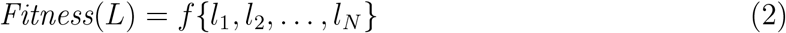

From the generated population, the %N population of Lions is considered as noma lions, and the rest is randomly divided into P Prides. The gender considered in the population remains constant throughout the optimization process. Out of P Prides %S (75%-90%) is considered as Female, the rest as male. In the case of Nomad, the ratio is vice versa %(1-S). In each searching stage, each position of the Lion is considered as the best position, and accordingly, the pride territory is determined, where the marked position in the pride territory is also considered as the best position of each pride.

#### 3.1.2. Hunting Phase

Lionesses hunt in groups to provide food for their pride, using specific strategies to encircle and catch prey. Stander et al. [44] classified lions into seven stalking roles, grouped into Left Wing, Centre, and Right-Wing positions, and each lioness corrects its position based on its own position during hunting. As the lions attack the prey from the opposite direction and cover almost a 360-degree attacks, to achieve this architecture, the Opposition-Based Learning (OBL) technique is used to explain the mathematical explanation of this study. In the hunting phase, the lions are divided into three different categories according to their fitness value. Lions with high fitness attack from the center, where, as the other two groups attack from the left and right wings of the prey, as depicted in the figure below. To achieve this, a dummy Prey(P) is chosen for centre 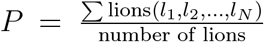. The hunting process involves randomly selecting hunters and attacking dummy prey, which will be established later based on the lion’s group. When a hunter improves its finesses, PREY(P) will flee, and a new location is achieved using Equation 3.

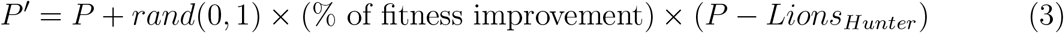

Where, P depicts the current_position of the prey, Lions_Hunter is the new position that attacks P. To present the encircling attack, the new positions of the Lions_Hunter of the left and right wings are presented using the Equation 4.

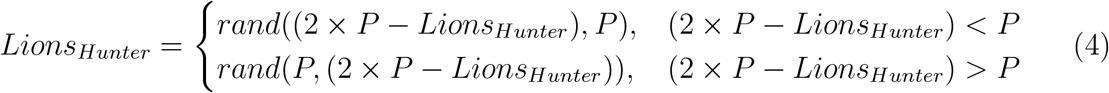

Where, P and Lions_Hunter present the current_position of P and *Lions*_*Hunter*_. The new position of the *Lions*_*Hunter*^*i*^ who attacks from the center is presented using Equation 5..

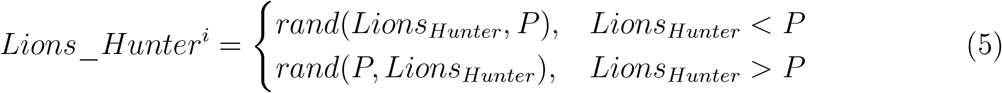

The proposed hunting method offers certain advantages to achieve improved solutions. An exceptional aspect of this tactic is that it creates a circular perimeter around the prey, enabling hunters to approach from several directions. Secondly, this method offers a means for solutions to evade local optima, as certain hunters employ opposing positions. The pseudocode of the overall mechanism is presented in algorithm 1.

##### Algorithm 1

Hunting Phase

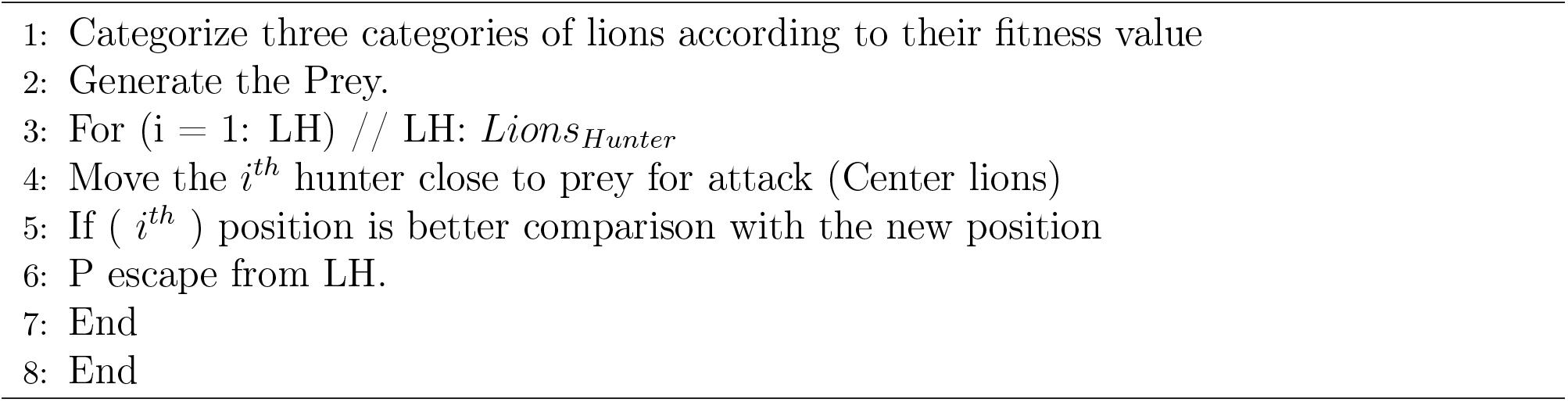

##### Position status of Lions (not involved in the hunting phase)

The position of the lions (Female Lion 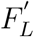 who are not involved in the hunting phase remained in the territory safely. The position of each member can be presented by Equation 6.

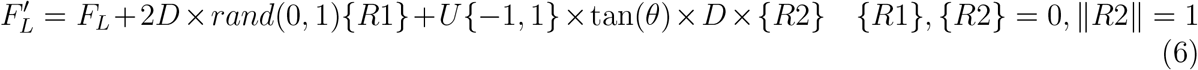

Here, the current and updated positions of the female lions are presented as *F*_*L*_ and *F*_*L*_*′*, respectively. *D* denotes the distance *F*_*L*_ from the pride’s territory (selected point) using the tournament selection approach. *R*1 is the vector presentation of the previous *F*_*L*_ and its direction to the selected point, whereas the vector *R*2 is perpendicular to vector *R*1. If the position of the lion improves until the last iteration (t), the success of the lion (i) can be updated in the group using Equation 7. where, 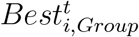 depicts the best position of the lion until iteration (t).

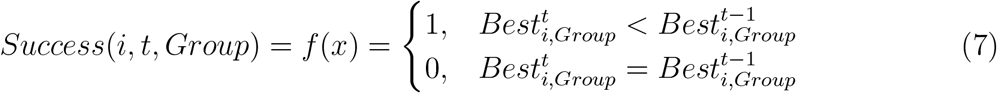

If the value of *Success*(*i, t, Group*) is high, it indicates that the lion’s position is far from the optimal point; otherwise, lions swing near the optimal point without any significant improvement in position. So, the count of lions in the pride (*j*) can be expressed using Equation 8, which improves its fitness value in each iteration. It played an important role in the decision on the size of the tournament.

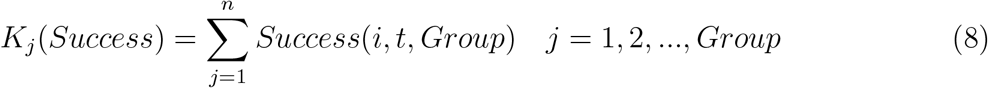

Here, n presents the count of lions in a pride. The value of *K*_*j*_(*Success*) is inversely proportional to tournament size. So, the tournament size is calculated using Equation 9. The position status of the lions is presented in Algorithm 2

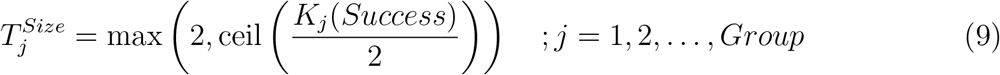

###### Algorithm 2

Position Status of Lions

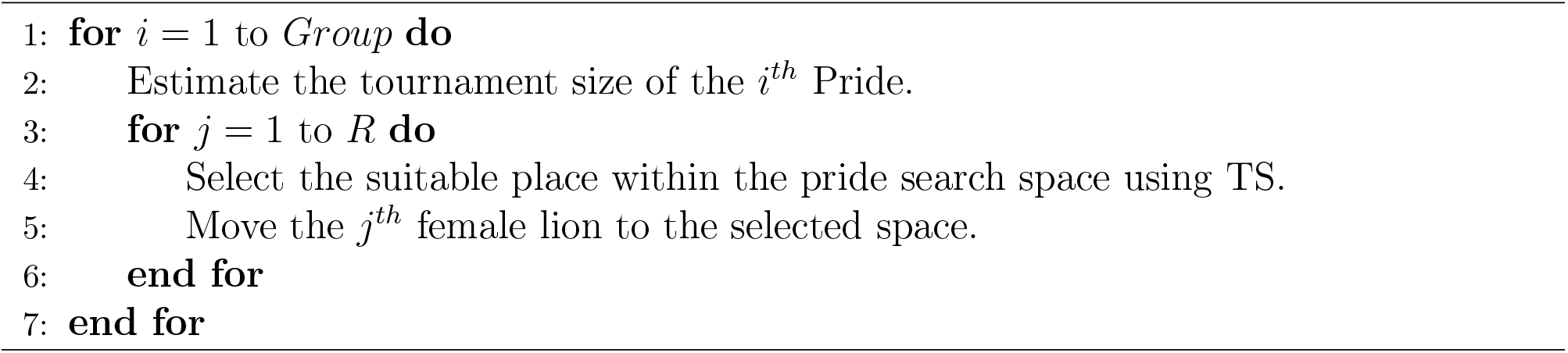

### 3.2. Roaming

Each male lion roams the territory of its pride for specific reasons. To replicate the behavior of resident males, the pride territories are randomly selected and subsequently visited by that lion. During traversing, if a resident lion encounters a new position that surpasses his current optimal position, update his best-visited solution. Algorithms 3 and 4 present the behavior of the male and the Nomad lion, respectively.

#### Algorithm 3

Territory Exploration by Remaining Male Lions

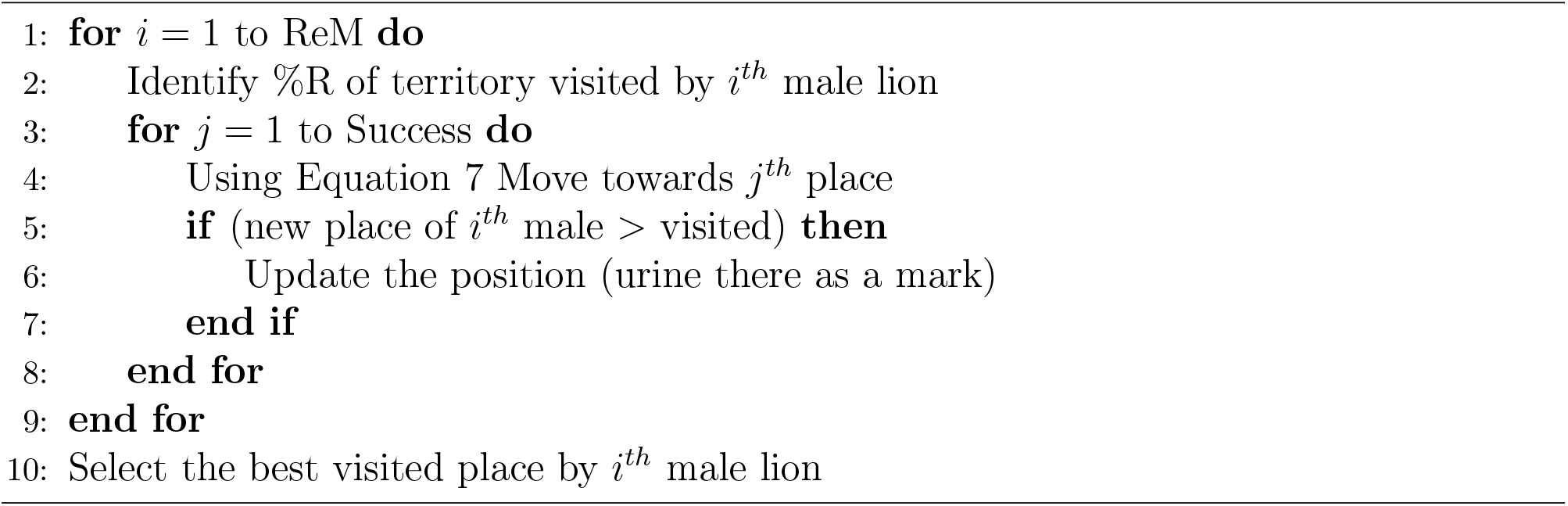

#### Algorithm 4

Movement of Nomadic Lions

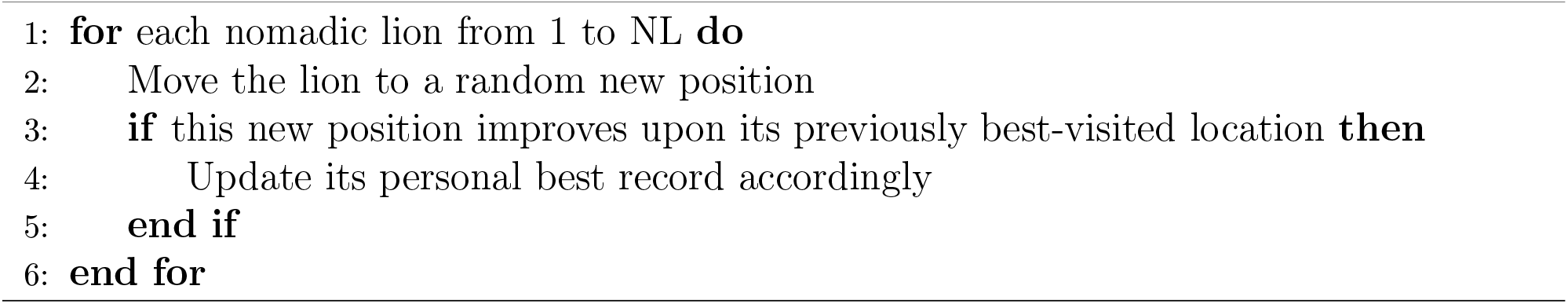

The movement of nomad lions and their neighbors in the search space in a random manner helps to avoid the trap in the local optima. The updated position of the nomad lion can be expressed using Equations 10 and 11.

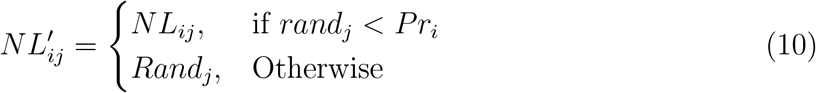

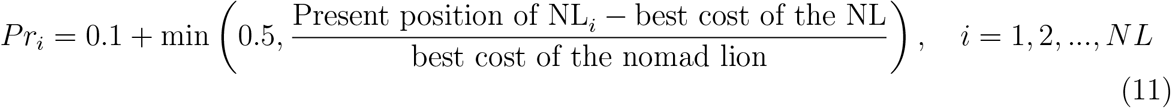

#### 3.2.1. Mating Mechanism

Mating is crucial for the survival of lions and the possibility of sharing information among members. In each pride, female lions tend to mate with one or more males. These men are randomly selected from the same pregnant female to generate children. Nomad female lions can only mate with one of the randomly selected males. The mating operoris produce two offspring through a linear combination of parents. The following equations 12 and 13 produce new cubs after selecting the female lion and male(s) format:

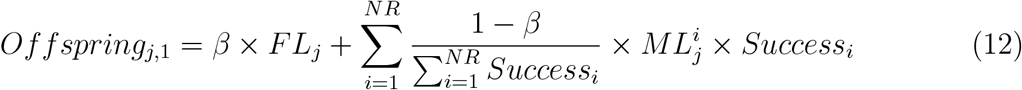

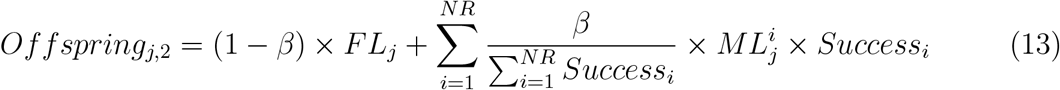

Here, *j* depicts the dimensions, the value of *Success*_*i*_ is 1 if selected for mating, else 0, NR depicts the number of ML (male lions) present, *β* is the random value whose mean is and standard deviation is 0.1. Of the two upspring, one is chosen for ML and the other for FL. After mating, the produced cub will inherit the features of both offsprings.

##### Defense

The existence of a lion (from inside the pride or a nomad one) in the pride depends on the fitness function or the strength. The winner is allowed to stay in the pride with the female lions. The defense operator is chosen in LO with the considerations depicted in Algorithm 5.

- Winner vs. existing male lions.
- Winner vs. nomad male lions.

###### Algorithm 5

Defense against nomad males

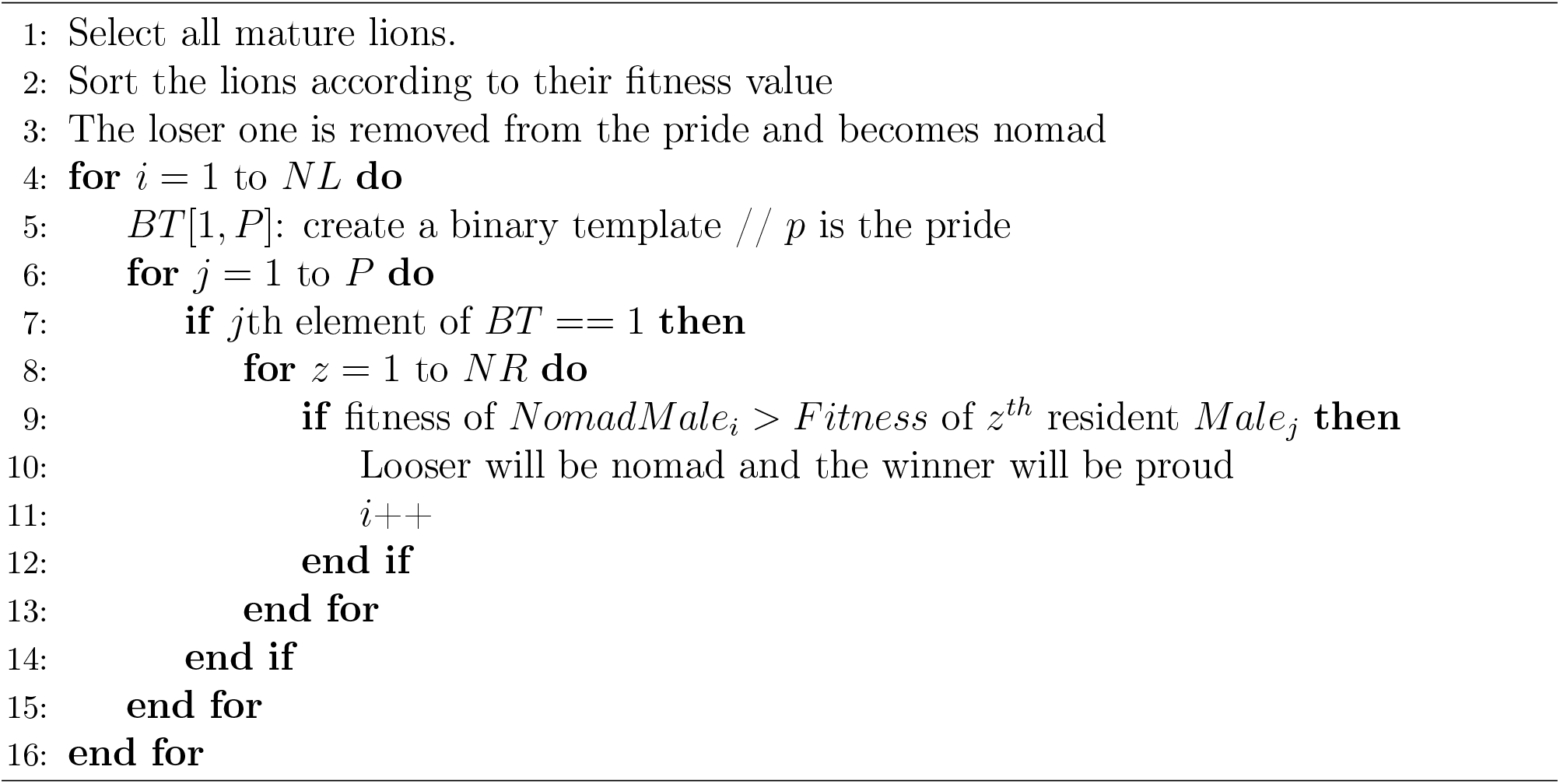

#### 3.2.2. Migration

Inspired by the migratory and switch lifestyle of lions in nature, when a lion migrates from one pride to another or changes its lifestyle, causing resident females to become nomads and vice versa, it increases the diversity of the target pride by its position in the preceding pride. On the other hand, the bridge for information sharing is built by the migration and switch lifestyle of Lions. The detailed description of LO is presented in Algorithm 6.

### 3.3. Binary Lion Optimization Algorithm (BLO) for Feature Selection

Feature selection is to identify the most useful features from a dataset, hence lowering dimensionality and improving the computational economy and predictive accuracy of machine learning models. Formulated as a binary optimization problem, this work represents each feature as a binary variable—chosen (1) or not selected (0). Operating in a continuous search space, the original Lion Optimization Algorithm (LO) has to be modified to operate in a binary domain to solve feature selection concerns. This adaptation transforms LOA into a binary variation exploring the binary solution space to find the best feature subset. Effective navigation of this discrete search space using binary implementations of LO, including the Binary Lion Optimization Algorithm (BLO), results in the choice of the most relevant characteristics. This, therefore, helps to increase model performance concerning computing cost, generalization, and accuracy. A detailed study of the proposed BLO is presented in Algorithm 7.

#### Algorithm 6

Lion Optimization Algorithm

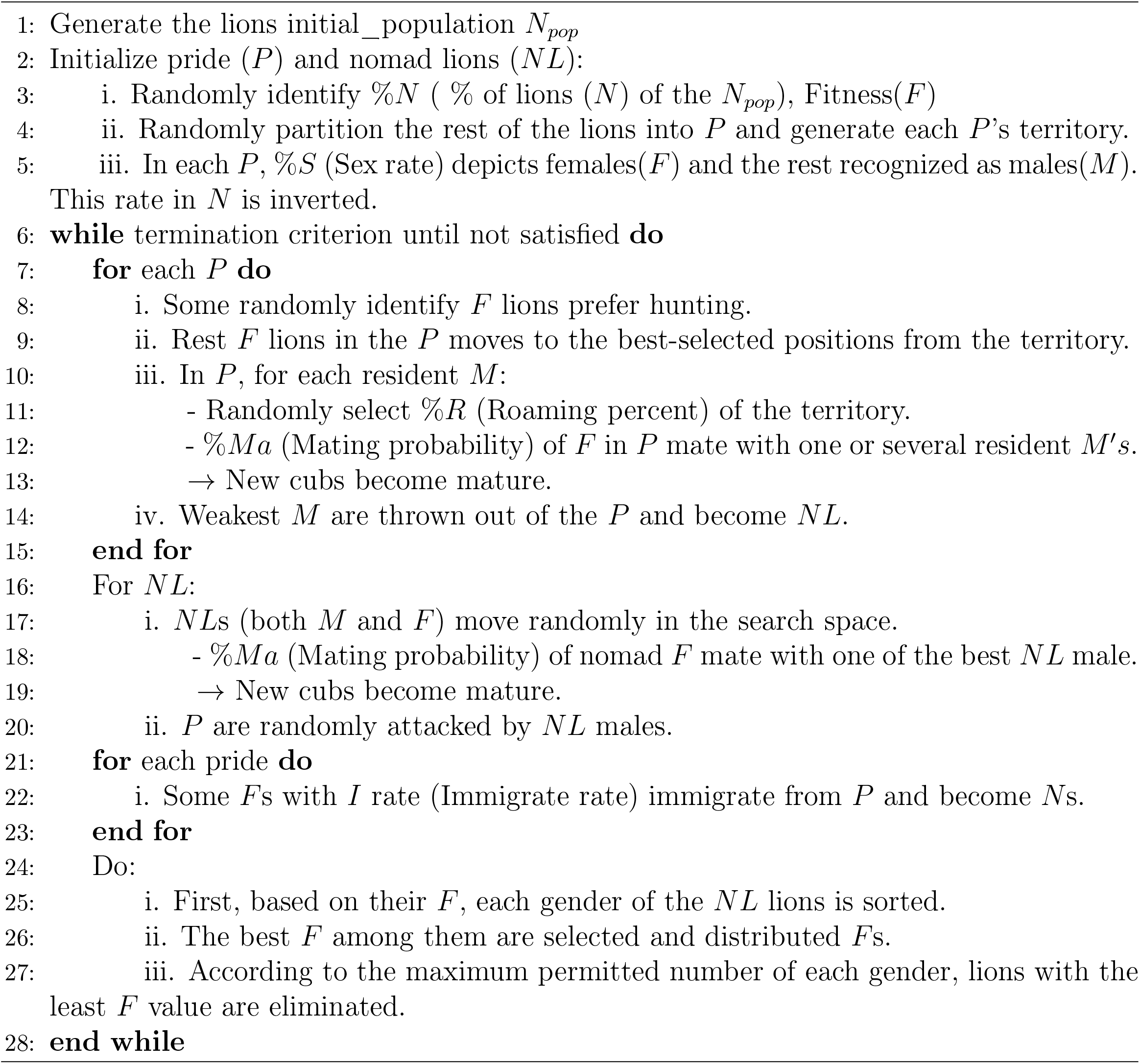

#### Algorithm 7

Binary Lion Optimization Algorithm

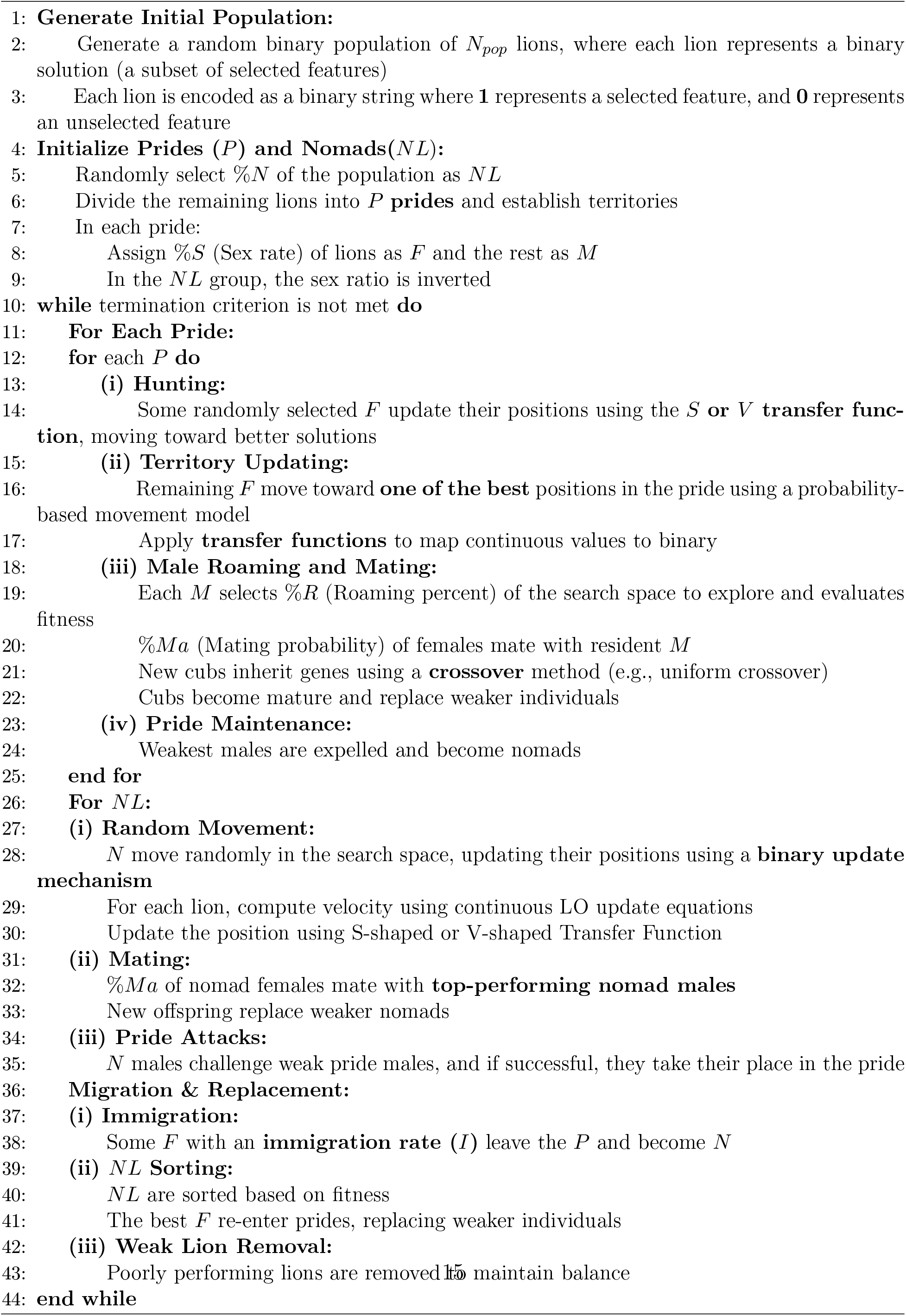

### 3.4. The mRMR filtering approach

The minimum Redundancy Maximum Relevance (mRMR) is one of the most prevalent feature selection (FS) methods in the literature. It uses an information-theoretic feature selection technique that selects features that have strong correlation and relevance to the target class (output), while reducing the intraclass feature redundancy. It uses the Mutual Information Difference (MID) objective function to evaluate genes in the search area. The measure of *MID* describes the amount of information a gene contributes to an organism’s phenotype, or in other words, how “useful” the gene is for the organism in the context; on the other hand, how much *MID* describes the redundancy other genes have as a correlate with other genes. Consider a microarray gene expression matrix 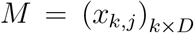, where *x*_*k,j*_ is the expression level of the gene *j* in sample *k*. The matrix contains *K* samples and *D* genes. Let the expression values of gene j for all samples be *x*_*j*_ = *x*_1*j*_, *x*_2*j*_,, *x*_*kj*_. The indexed set of genes is expressed as *G* = 1, 2, … ., *D*. The mRMR algorithm takes the first step of the selection process by selecting the gene with the greatest mutual information concerning the class label *𝓁*, which is included in the subset *Z* ⊂ *G* [24]. Equations 14 and 15 represent the formulas to calculate the MID and MI.

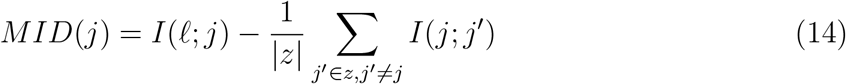

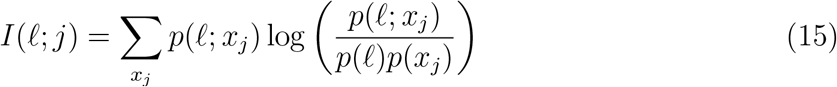

Here, *I*(*𝓁*; *j*) depicts the MI between the class level (*𝓁*) and the gene (*j*) while *I*(*j*; *j′* ) presents the redundancy of the gene (*j*) compared to other genes in subset Z.

### 3.5. Fitness function

The fitness function considers not only the accuracy of the classification, but also the chosen features, too. Its objective is to achieve the highest possible classification accuracy for a minimum subset of characteristics. For this purpose, the fitness function adopted to assess particular solutions is described in Eq. 16.

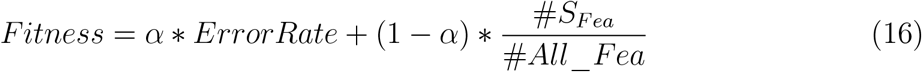

In this case, ErrorRate is the measure of the classification errors for the chosen features. It is estimated as the number of wrongly classified cases (by the 5-KNN classifier) over the total number of classifications, expressed as a ratio from 0 to 1. Because classification accuracy is the proportion of correctly resolved instances, a low value of Error Rate suggests that the system’s accuracy is high. In addition, #*S*_*F ea*_ refers to the features that were selected in the experiment, while #*All*_*F ea*_ refers to the class of features available in the data set. The parameter a adjusts the degree to which classification quality and the size of the subset of characteristics are balanced. In our experiments, this value was set at 0.9. The classification error is assessed as the ratio of misclassified predictions to the sum of all predictions given, as a number between 0 and 1.

### 3.6. The proposed mRMR-BLO algorithm for Feature Selection

This section presents a novel feature selection (FS) approach that integrates the mRMR filtering technique with two variants of the Lion Optimization (LO) algorithm using two different transfer functions. Initially, the mRMR method is employed to eliminate the least important genes from the dataset. Then, as illustrated in Fig. 2, the proposed LO algorithm, utilizing S and V transfer functions, is used to identify the most relevant gene subset from the remaining genes. A detailed explanation of this method is provided in the following subsections.

**Figure 2:**
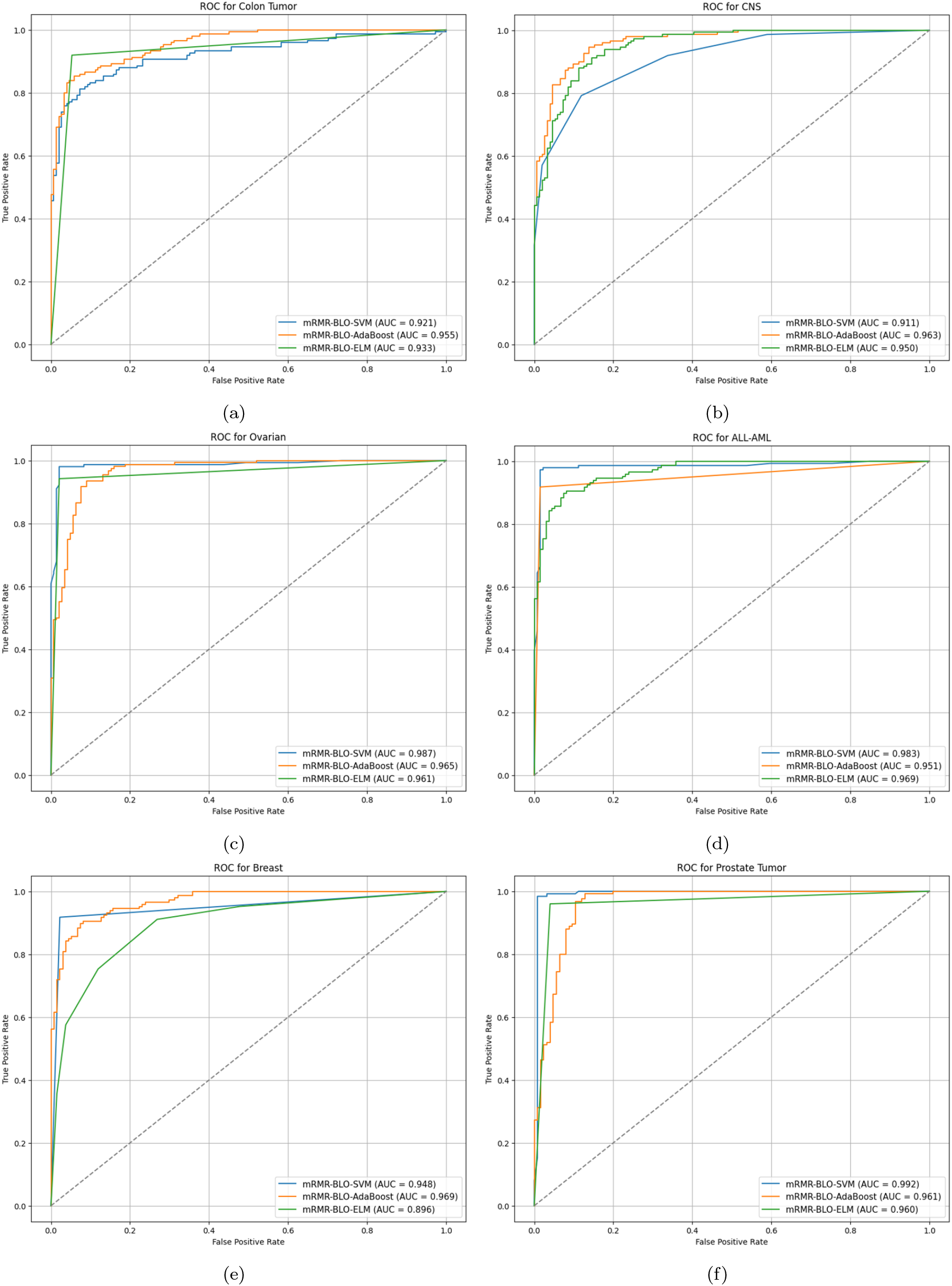
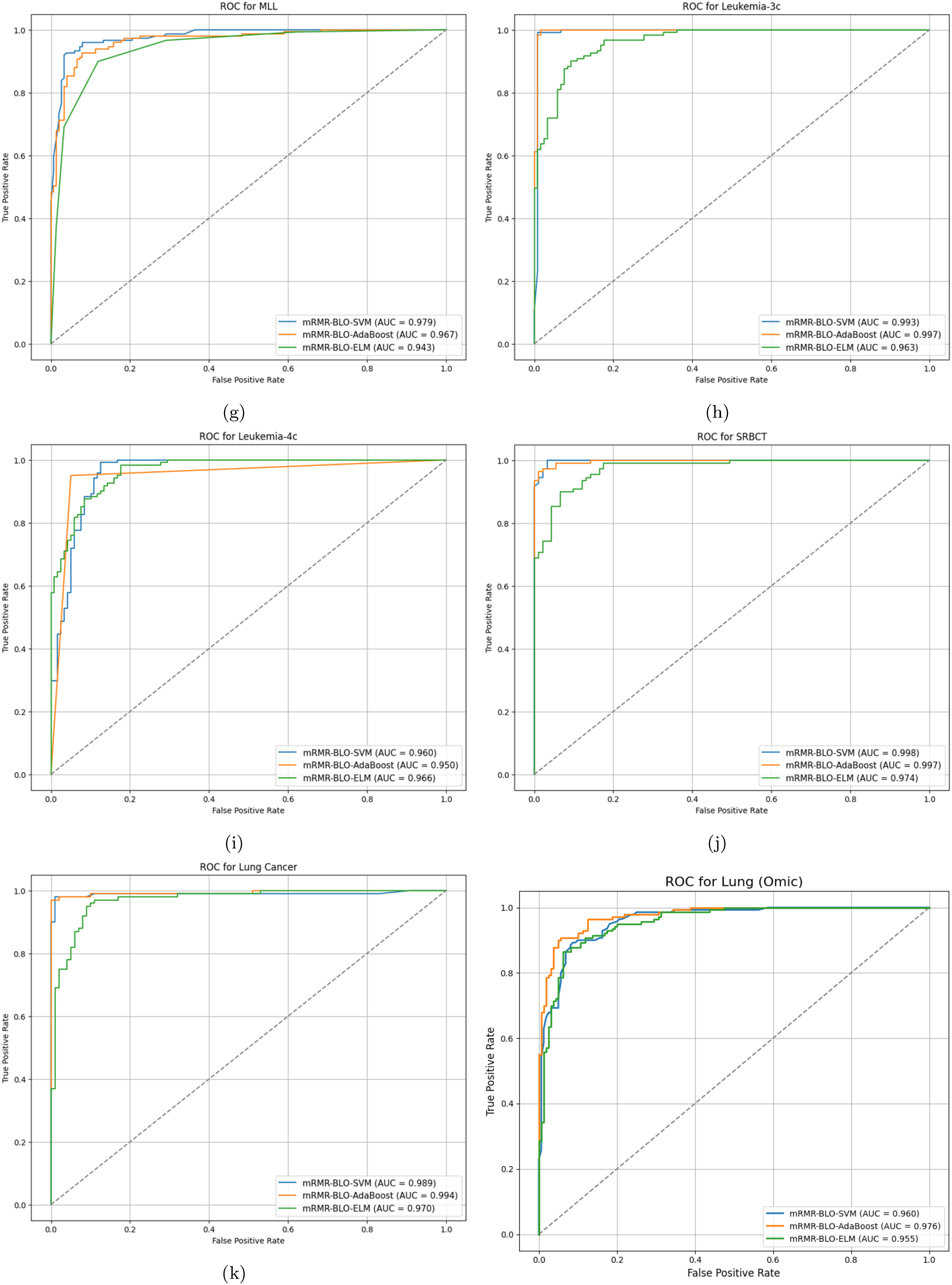
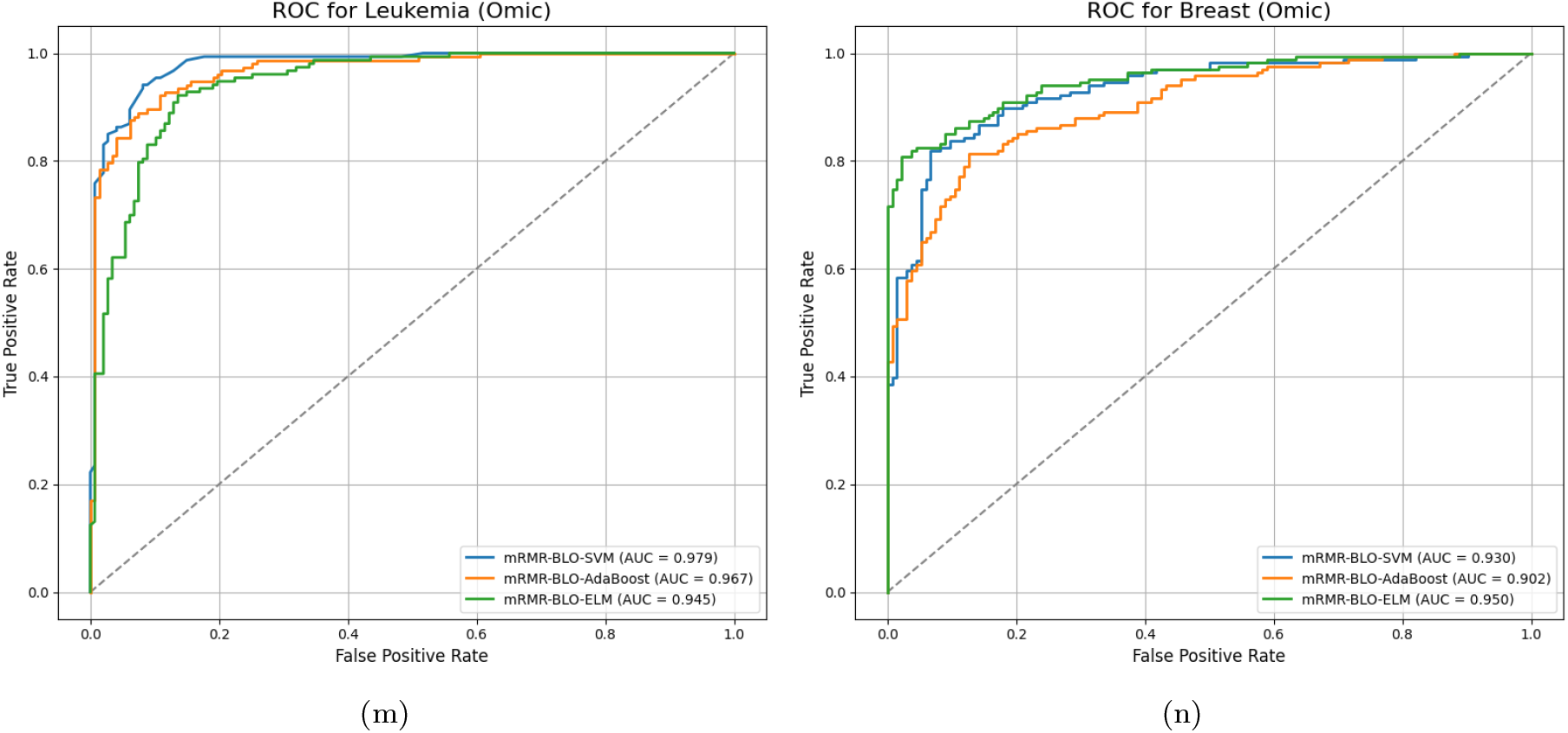
ROC analysis for (a) Colon Tumor (b) CNS (c) Ovarian (d) ALL-AML (e) Breast Cancer (f) Prostate Tumor (g) MLL (h) Leukemia-3c (i) Leukemia-4c (j) SRBCT (K) Lung Cancer (l) Lung (Omic) Cancer (m) Leukemia (Omic) Cancer (n) Breast (Omic) Cancer dataset

#### 3.6.1. Feature Preprocessing Using mRMR

The mRMR filtering method is applied as a preprocessing step to refine the input data for the proposed Binary Lion Optimization (BLO) by removing noisy genes. This method selects the M highest-ranked genes based on their correlation with each other and the class label. The chosen genes serve as the input for initializing the population in the BLO wrapper-based FS approach, making them more effective for cancer classification. Besides enhancing classification accuracy, the filtering process significantly reduces computational complexity. This is particularly important because, in wrapper-based approaches, the number of possible gene subsets increases exponentially as the number of genes grows, making an exhaustive search computationally expensive.

#### 3.6.2. Feature Selection Using the BLO Wrapper Approach

The features selected during the preprocessing stage are passed on to the proposed BLO wrapper approach to determine the optimal gene subset. At this stage, the M top-ranked genes from mRMR are further refined to obtain the smallest subset of informative genes with the highest fitness value for cancer classification. Gene selection (GS) is a binary optimization problem, where solutions consist of binary values (0 and 1). A candidate solution X is represented as a binary string of length D, denoted *X* = { *g*_1_, *g*_2_, …, *g*_*D*_ }, where *D* is the dimension of the problem, and *g*_*j*_ represents the gene *j*^*th*^ (*j* = { 1, 2, …, *D* } ). In this binary representation, a gene *g*_*j*_ is retained if its value is 1 and discarded if it is 0. Thus, only genes encoded as 1 are considered for evaluation. Since LO (Lion Optimizer) operates in a continuous search space, whereas the GS problem exists in a discrete (binary) space, the continuous values must be converted into binary form before applying LO to GS. This requires modifying LO to accommodate binary optimization, ensuring that each element in a solution is either 0 or 1. This transformation is achieved through a transfer function (TF), which maps continuous values to binary values. There are various types of transfer functions available for this purpose.

## 4. Experimental results and performance comparison

The algorithms considered were evaluated using 11 publicly available microarray gene expression datasets for different diseases https://data.mendeley.com/datasets/fhx5zgx2zj/ 1 and 3 Omic cancer datasets from https://sbcb.inf.ufrgs.br/cumida#datasets. The details of this data set are detailed in Table 1, which consists of the title of the data set, sample size, number of genes, number of classes, and diagnostic task. The datasets containing thousands of genes fitted both binary and multiclass classifications. The datasets are high-dimensional data with dimensionality ranging from 2000 to 15154, while the number of samples (patients) is relatively small. The performance of these proposed algorithms was evaluated using an SVM classifier with 10-fold cross-validation on a single dataset, while the Leave-One-Out cross-validation (LOOCV) method provided the final evaluation of the proposed methodology with the selected genes. In addition, a linear kernel was used in the SVM for the classification problem. To guarantee fairness in the experiment, each technique was performed 10 times per dataset, and the final numbers were returned as the average. Table 2 contains the configuration values for the proposed methods. The listed values were established after some exploratory tests and known studies on LO.

**Table 1:** Exploration of Datasets.

| **Dataset Name** | **#Samples** | **#Genes** | **#Classes** |
| --- | --- | --- | --- |
| Colon Tumor | 62 | 2000 | 2 |
| CNS | 60 | 7129 | 2 |
| Ovarian | 253 | 15154 | 2 |
| ALL-AML | 72 | 7129 | 2 |
| Breast Cancer | 97 | 24481 | 2 |
| Prostate Tumor | 102 | 10509 | 2 |
| MLL | 72 | 12582 | 3 |
| Leukemia-3c | 72 | 7129 | 3 |
| Leukemia-4c | 72 | 7129 | 4 |
| SRBCT | 83 | 2308 | 4 |
| Lung Cancer | 203 | 12600 | 5 |
| Breast(Omic) | 66 | 22278 | 3 |
| Lung(Omic) | 90 | 54676 | 2 |
| Leukemia(Omic) | 52 | 22284 | 2 |

**Table 2:**
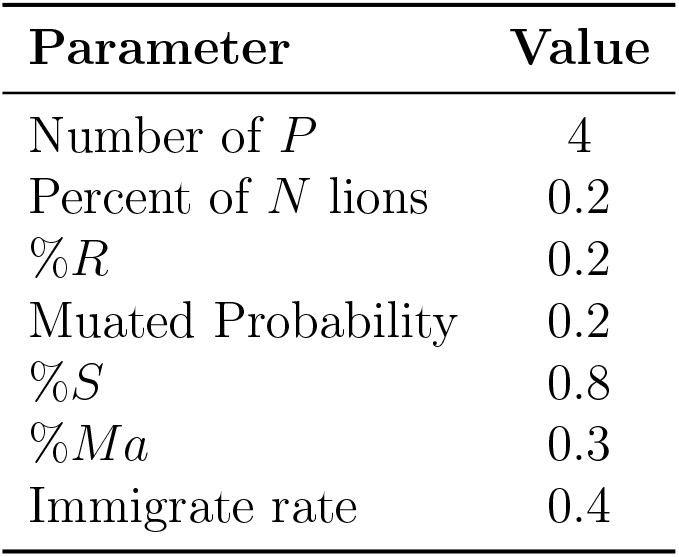
Binary Lion Optimization Algorithm Parameters.

Table 3 shows the performance of various filter approaches in selecting features from different microarray datasets. The effectiveness of feature selection techniques for the Minimum Redundancy (mRMR, IG, Chi-square, and ReliefF methods has been analyzed in different binary and multiclass datasets. The findings suggest that mRMR is superior to all other filter methods in all empirical cases, consistent with the existing literature on feature selection. In binary classification, mRMR dominates as the top performer in five of six datasets. mRMR has considerable margins over IG, chi-square, and ReliefF in colon tumor (85.48%), CNS (75%), ovarian (96.12%), breast cancer (75.25%), and prostate tumor (94.11%). The ALL-AML data set is an exception, in which mRMR and IG were marked with the highest accuracy of 98.61%. These figures portray how mRMR’s inbuilt feature redundancy elimination fostered performance excellence and drove mRMR dominance in binary classification tasks. For multiclass classification tasks, mRMR stays on top in three out of five datasets: MLL (97.22%), SRBCT (92.78%), and Lung Cancer (96.55%). In the Leukemia-3c dataset, mRMR is marginally surpassed by IG, and for Leukemia-4c, mRMR, IG, and Chi-square lockstep achieve the same accuracy. Although accuracy, these alterations leave mRMR with an overall lead in multiclass classification. The findings uphold mRMR as the most efficient filter-based feature selection technique for microarray datasets. It’s ability to balance the relevance and redundancy for selecting the most informative features from the entire feature set for which it may be considered as the preferable filter approach in contrast to other filter approaches considered.

**Table 3:** Classification Accuracy of Feature Selection Methods on Cancer Datasets(%)

| Class | Dataset | Metric | mRMR | IG | Chi-square | Relieff |
| --- | --- | --- | --- | --- | --- | --- |
| Binary Class | Colon Tumor | Accuracy | 85.48 | 77.41 | 79.03 | 80.64 |
|  | CNS |  | 75.00 | 63.33 | 73.33 | 66.66 |
|  | Ovarian |  | 96.12 | 90.17 | 89.87 | 87.12 |
|  | ALL-AML |  | 98.61 | 98.61 | 97.22 | 97.22 |
|  | Breast Cancer |  | 75.25 | 74.22 | 74.22 | 77.31 |
|  | Prostate Tumor |  | 94.11 | 88.23 | 86.27 | 93.13 |
|  | Lung(Omic) |  | 89.54 | 79.85 | 86.93 | 84.44 |
|  | Leukemia(Omic) |  | 85.79 | 79.88 | 86.12 | 79.89 |
| Multi Class | MLL | Accuracy | 97.22 | 95.83 | 97.22 | 97.22 |
|  | Leukemia-3c |  | 95.83 | 97.22 | 94.44 | 94.44 |
|  | Leukemia-4c |  | 94.44 | 94.44 | 94.44 | 87.50 |
|  | SRBCT |  | 92.78 | 86.78 | 85.10 | 82.87 |
|  | Lung Cancer |  | 96.55 | 93.59 | 92.11 | 91.62 |
|  | Breast (Omic) |  | 78.84 | 82.96 | 81.19 | 79.11 |

### 4.1. Performance Evaluation of BLO with Different Classifiers

Table 4 shows the performance of BLO with different classifiers such as SVM, ELM, and AdaBoost. The outcomes indicate that the BLO-SVM routinely performs well on several datasets, often achieving high accuracy. BLO-AdaBoost also demonstrates competitive performance, particularly for ovarian and colon cancer datasets.

**Table 4:** Performance metrics for different models on various datasets (%).

| Datasets | Model | Accuracy | MCR | PRE | REC | F1-S | SPE | FNR | FPR | MCC |
| --- | --- | --- | --- | --- | --- | --- | --- | --- | --- | --- |
| Colon Tumor | BLO-SVM | 87.50 | 12.50 | 76.92 | 86.96 | 81.63 | 87.76 | 13.04 | 12.24 | 72.52 |
|  | BLO-AdaBoost | 88.89 | 11.11 | 88.00 | 81.48 | 84.62 | 93.33 | 18.52 | 6.67 | 76.08 |
|  | BLO-ELM | 88.57 | 11.43 | 80.65 | 92.59 | 86.21 | 86.05 | 7.41 | 13.95 | 77.06 |
| CNS | BLO-SVM | 88.10 | 11.90 | 85.71 | 90.00 | 850 | 86.36 | 10.00 | 13.64 | 76.28 |
|  | BLO-AdaBoost | 89.47 | 10.53 | 91.84 | 88.24 | 90.00 | 90.91 | 11.76 | 9.09 | 78.97 |
|  | BLO-ELM | 86.75 | 13.25 | 83.33 | 89.74 | 86.42 | 84.09 | 10.26 | 15.91 | 73.71 |
| Ovarian | BLO-SVM | 94.12 | 5.88 | 97.78 | 91.67 | 94.62 | 97.30 | 8.33 | 2.70 | 88.37 |
|  | BLO-AdaBoost | 95.18 | 4.82 | 95.74 | 95.74 | 95.74 | 94.44 | 4.26 | 5.56 | 90.19 |
|  | BLO-ELM | 86.75 | 13.25 | 83.33 | 89.74 | 86.42 | 84.09 | 10.26 | 15.91 | 73.71 |
| ALL-AML | BLO-SVM | 97.65 | 2.35 | 97.92 | 97.92 | 97.92 | 97.30 | 2.08 | 2.70 | 95.21 |
|  | BLO-AdaBoost | 97.59 | 2.41 | 97.92 | 97.92 | 97.92 | 97.14 | 2.08 | 2.86 | 95.06 |
|  | BLO-ELM | 90.36 | 9.64 | 90.48 | 90.48 | 90.48 | 90.24 | 9.52 | 9.76 | 80.72 |
| Breast | BLO-SVM | 854 | 12.16 | 77.78 | 87.50 | 82.35 | 88.00 | 12.50 | 12.00 | 73.42 |
|  | BLO-AdaBoost | 88.89 | 11.11 | 87.50 | 80.77 | 84.00 | 93.48 | 19.23 | 6.52 | 75.65 |
|  | BLO-ELM | 90.14 | 9.86 | 83.87 | 92.86 | 88.14 | 88.37 | 7.14 | 11.63 | 80.04 |
| Prostate Tumor | BLO-SVM | 97.59 | 2.41 | 98.08 | 98.08 | 98.08 | 96.77 | 1.92 | 3.23 | 94.85 |
|  | BLO-AdaBoost | 95.18 | 4.82 | 95.74 | 95.74 | 95.74 | 94.44 | 4.26 | 5.56 | 90.19 |
|  | BLO-ELM | 95.06 | 4.94 | 95.92 | 95.92 | 95.92 | 93.75 | 4.08 | 6.25 | 89.67 |
| MLL | BLO-SVM | 92.94 | 7.06 | 95.56 | 91.49 | 93.48 | 94.74 | 8.51 | 5.26 | 85.89 |
|  | BLO-AdaBoost | 93.83 | 6.17 | 93.33 | 95.45 | 94.38 | 91.89 | 4.55 | 8.11 | 87.56 |
|  | BLO-ELM | 87.21 | 12.79 | 85.71 | 87.80 | 86.75 | 86.67 | 12.2 | 13.33 | 74.41 |
| Leukemia-3c | BLO-SVM | 94.12 | 5.88 | 95.56 | 93.48 | 94.51 | 94.87 | 6.52 | 5.13 | 88.2 |
|  | BLO-AdaBoost | 93.75 | 6.25 | 95.35 | 93.18 | 94.25 | 94.44 | 6.82 | 5.56 | 87.43 |
|  | BLO-ELM | 91.57 | 8.43 | 94.74 | 87.80 | 91.14 | 95.24 | 12.2 | 4.76 | 83.33 |
| Leukemia-4c | BLO-SVM | 95.29 | 4.71 | 95.65 | 95.65 | 95.65 | 94.87 | 4.35 | 5.13 | 90.52 |
|  | BLO-AdaBoost | 95.06 | 4.94 | 97.73 | 93.48 | 95.56 | 97.14 | 6.52 | 2.86 | 90.12 |
|  | BLO-ELM | 93.90 | 6.1 | 94.87 | 92.50 | 93.67 | 95.24 | 7.5 | 4.76 | 852 |
| SRBCT | BLO-SVM | 97.62 | 2.38 | 97.87 | 97.87 | 97.87 | 97.30 | 2.13 | 2.7 | 95.17 |
|  | BLO-AdaBoost | 97.56 | 2.44 | 97.87 | 97.87 | 97.87 | 97.14 | 2.13 | 2.86 | 95.02 |
|  | BLO-ELM | 90.91 | 9.09 | 91.49 | 91.49 | 91.49 | 90.24 | 8.51 | 9.76 | 81.73 |
| Lung Cancer | BLO-SVM | 86.36 | 13.64 | 70.00 | 82.35 | 75.68 | 87.76 | 17.65 | 12.24 | 66.71 |
|  | BLO-AdaBoost | 97.59 | 2.41 | 97.92 | 97.92 | 97.92 | 97.14 | 2.08 | 2.86 | 95.06 |
|  | BLO-ELM | 95.24 | 4.76 | 95.83 | 95.83 | 95.83 | 94.44 | 4.17 | 5.56 | 90.28 |
| Lung(Omic) | BLO-SVM | 92.22 | 7.78 | 97.22 | 85.37 | 90.91 | 97.96 | 14.63 | 2.04 | 84.71 |
|  | BLO-AdaBoost | 95.56 | 4.44 | 97.44 | 92.68 | 95.00 | 97.96 | 7.32 | 2.04 | 91.10 |
|  | BLO-ELM | 93.33 | 6.67 | 93.94 | 88.57 | 91.18 | 96.36 | 11.43 | 3.64 | 85.92 |
| Leukemia(Omic) | BLO-SVM | 90.38 | 9.62 | 90.00 | 93.10 | 91.53 | 86.96 | 6.90 | 13.04 | 80.48 |
|  | BLO-AdaBoost | 94.23 | 5.77 | 95.45 | 91.30 | 93.33 | 96.55 | 8.70 | 3.45 | 88.32 |
|  | BLO-ELM | 92.31 | 7.69 | 92.59 | 92.59 | 92.59 | 92.00 | 7.41 | 8.00 | 84.59 |
| Breast (Omic) | BLO-SVM | 86.36 | 13.64 | 70.00 | 82.35 | 75.68 | 87.76 | 17.65 | 12.24 | 66.71 |
|  | BLO-AdaBoost | 87.88 | 12.12 | 84.21 | 76.19 | 80.00 | 93.33 | 23.81 | 6.67 | 71.52 |
|  | BLO-ELM | 89.39 | 10.61 | 81.48 | 91.67 | 86.27 | 88.10 | 8.33 | 11.90 | 78.04 |

- With an accuracy of 88.89% for the Colon Tumour dataset, BLO-AdaBoost exceeded BLO-SVM and BLO-ELM by 1.39% and 0.32%, respectively. In addition, showing dependability in classification, BLO-AdaBoost obtained a high specificity of 93.33% and an MCC score of 76.08%. Moreover, it was suggested that BLO-AdaBoost offered balanced categorization and thus was appropriate for the dataset, as were the acquired precision and recall values.
- With an accuracy of 89.47%, which was an increase of 1.37% over BLO-SVM 88.10% and 2.72% over BLO-ELM 86.75%, BLO-AdaBoost emerged once more in the CNS dataset. Moreover, it attained great accuracy and recall values—as shown by an MCC of 78.97%, which is the best categorization among all. The preserved sensitivity and specificity balance of BLO-AdaBoost made it practical for the dataset.
- BLO-AdaBoost once more reached the best accuracy at 95.18% for the ovarian dataset. BLO-SVM and BLO-ELM were exceeded in this dataset by BLO-AdaBoost by 1.06% and 8.43%, respectively. High recall and specificity values, together with an MCC score of 90.19%, validated BLO-AdaBoost’s classification power. Enough positive and negative class differences showed that the model worked effectively at discriminating, with low false classification rates.
- With a slight 0.06% gain from BLO-AdaBoost (97.59%) and a notable 7.29% rise from BLO-ELM (90.36%), BLO-SVM attained the highest accuracy in the ALL-AML dataset at 97.65%. Indicating great classification power, it kept great specificity at 97.30% and an MCC of 95.21%. The great accuracy and recall figures show that BLO-SVM identified positive and negative situations really brilliantly.
- With an accuracy of 90.14%, BLO-ELM surpassed other models for the Breast dataset by 2.30% and BLO-SVM (854%) by 1.25%. Additionally, demonstrating great sensitivity and specificity, it turned out to be a useful classifier for the dataset. The MCC of 80.04% backs up the assertion of exceptional categorization capacity.
- With an accuracy of 97.59%, which outperformed BLO-AdaBoost (95.18%) and BLO-ELM (95.06%) by 2.41% and 2.53%, respectively, BLO-SVM had the best performance in the Prostate Tumour dataset. It also presented an excellent MCC of 94.85%, therefore demonstrating good classification accuracy. The dependability of BLO-SVM’s forecasts is shown by low false negative and false positive rates. With 93.83% accuracy, BLO-AdaBoost excelled with an improvement of 0.89% on BLO-SVM’s 92.94% and 6.62% on BLO-ELM’s 87.21%. Strong in illness categorization, this classifier also attained a high recall rate of 95.45%. With an MCC score of 87.56%, it keeps a consistent classification capacity.
- The best performance was shown in the Leukemia-3c dataset by BLO-SVM, whose accuracy score was 94.12%, outperforming BLO-AdaBoost with 93.75% and BLO-ELM with 91.57% by 0.37% and 2.55%, respectively. MCC score of 88.20% confirmed its better classification power. Precision implies that BLO-SVM was able to discriminate between various subtypes of leukemia and identify them precisely without merging.
- With a score of 95.29%, BLO-SVM with 0.23% and 1.39% higher than BLO-AdaBoost and BLO-ELM, with 95.06% and 93.90% respectively, attained the greatest accuracy for the dataset Leukemia-4c. Its MCC of 90.52% confirmed even more the dependability and accuracy of the classifier. The performance of the classifier revealed balanced sensitivity and specificity.
- In the SRBCT dataset, where an accuracy of 97.62 was recorded, BLO-SVM displayed the best performance with a 0.06% gain over BLO-AdaBoost and 6.71% over BLO-ELM, producing respectively a previously recorded 97.56% and 90.91%. It also attained a 97.30% specificity, further boosting classification accuracy. The low disparity between BLO-SVM and BLO-AdaBoost shows that the two models attained comparable performance.
- With a maximum accuracy of 97.65%, BLO-SVM outperformed BLO-AdaBoost (97.59%) by 0.06% and BLO-ELM (95.24%) by 2.41% on the Lung Cancer dataset. Furthermore, its great performance is supported by the MCC score of 95.21%. Its low rates of false positive and false negative errors imply that BLO-SVM performed highly for lung cancer detection.
- Using the Lung Cancer Omic dataset, the BLO-AdaBoost shows the highest accuracy of 95.56%. While BLO-ELM shows an accuracy of 93.33%, followed by BLO-SVM of 92.22%.
- With the Leukemia Omic dataset, the BLO-AdaBoost model outperforms the other two models with an accuracy of 94.23%.
- Considering the Breast Cancer Omic dataset, the BLO-ELM shows an accuracy level of 89.39%, followed by BLO-AdaBoost and BLO-SVM with accuracy of 87.88% and 86.36%, respectively.

### 4.2. Performance Evaluation of Proposed Model

Table 5 shows the performance of the proposed model with different datasets.

**Table 5:** Performance metrics for different models on various datasets (%).

| Datasets | Model | Accuracy | MCR | PRE | REC | F1-S | SPE | FNR | FPR | MCC |
| --- | --- | --- | --- | --- | --- | --- | --- | --- | --- | --- |
| Colon Tumor | mRMR-BLO-SVM | 91.55 | 8.45 | 89.36 | 97.67 | 93.33 | 82.14 | 2.33 | 156 | 82.46 |
|  | mRMR-BLO-AdaBoost | 94.44 | 5.56 | 94.23 | 98.00 | 96.08 | 86.36 | 2.00 | 13.64 | 86.76 |
|  | mRMR-BLO-ELM | 92.96 | 7.04 | 94.55 | 96.30 | 95.41 | 82.35 | 3.70 | 17.65 | 80.33 |
| CNS | mRMR-BLO-SVM | 92.86 | 7.14 | 96.77 | 93.75 | 95.24 | 90.00 | 6.25 | 10.00 | 81.13 |
|  | mRMR-BLO-AdaBoost | 95.60 | 4.40 | 97.30 | 97.30 | 97.30 | 88.24 | 2.70 | 11.76 | 85.53 |
|  | mRMR-BLO-ELM | 95.65 | 4.35 | 97.30 | 97.30 | 97.30 | 88.89 | 2.70 | 11.11 | 86.19 |
| Ovarian | mRMR-BLO-SVM | 97.56 | 2.44 | 98.08 | 98.08 | 98.08 | 96.67 | 1.92 | 3.33 | 94.74 |
|  | mRMR-BLO-AdaBoost | 95.12 | 4.88 | 95.74 | 95.74 | 95.74 | 94.29 | 4.26 | 5.71 | 90.03 |
|  | mRMR-BLO-ELM | 96.20 | 3.80 | 97.92 | 95.92 | 96.91 | 96.67 | 4.08 | 3.33 | 92.02 |
| ALL-AML | mRMR-BLO-SVM | 97.53 | 2.47 | 98.08 | 98.08 | 98.08 | 96.55 | 1.92 | 3.45 | 94.63 |
|  | mRMR-BLO-AdaBoost | 95.12 | 4.88 | 95.92 | 95.92 | 95.92 | 93.94 | 4.08 | 6.06 | 89.86 |
|  | mRMR-BLO-ELM | 96.20 | 3.80 | 957 | 95.83 | 96.84 | 96.77 | 4.17 | 3.23 | 92.11 |
| Breast | mRMR-BLO-SVM | 94.05 | 5.95 | 97.78 | 91.67 | 94.62 | 97.22 | 8.33 | 2.78 | 88.20 |
|  | mRMR-BLO-AdaBoost | 95.12 | 4.88 | 95.74 | 95.74 | 95.74 | 94.29 | 4.26 | 5.71 | 90.03 |
|  | mRMR-BLO-ELM | 86.59 | 13.41 | 83.33 | 89.74 | 86.42 | 83.72 | 10.26 | 16.28 | 73.40 |
| Prostate Tumor | mRMR-BLO-SVM | 97.89 | 2.11 | 98.41 | 98.41 | 98.41 | 96.88 | 1.59 | 3.13 | 95.29 |
|  | mRMR-BLO-AdaBoost | 97.75 | 2.25 | 98.18 | 98.18 | 98.18 | 97.06 | 1.82 | 2.94 | 95.24 |
|  | mRMR-BLO-ELM | 95.35 | 4.65 | 96.08 | 96.08 | 96.08 | 94.29 | 3.92 | 5.71 | 90.36 |
| MLL | mRMR-BLO-SVM | 97.56 | 2.44 | 97.87 | 97.87 | 97.87 | 97.14 | 2.13 | 2.86 | 95.02 |
|  | mRMR-BLO-AdaBoost | 97.78 | 2.22 | 98.18 | 98.18 | 98.18 | 97.14 | 1.82 | 2.86 | 95.32 |
|  | mRMR-BLO-ELM | 95.40 | 4.60 | 95.35 | 95.35 | 95.35 | 95.45 | 4.65 | 4.55 | 90.80 |
| Leukemia-3c | mRMR-BLO-SVM | 97.59 | 2.41 | 97.96 | 97.96 | 97.96 | 97.06 | 2.04 | 2.94 | 95.02 |
|  | mRMR-BLO-AdaBoost | 98.06 | 1.94 | 98.53 | 98.53 | 98.53 | 97.14 | 1.47 | 2.86 | 95.67 |
|  | mRMR-BLO-ELM | 95.92 | 4.08 | 96.30 | 96.30 | 96.30 | 95.45 | 3.70 | 4.55 | 91.75 |
| Leukemia-4c | mRMR-BLO-SVM | 96.70 | 3.30 | 98.18 | 96.43 | 97.30 | 97.14 | 3.57 | 2.86 | 93.10 |
|  | mRMR-BLO-AdaBoost | 96.67 | 3.33 | 98.18 | 96.43 | 97.30 | 97.06 | 3.57 | 2.94 | 92.98 |
|  | mRMR-BLO-ELM | 94.32 | 5.68 | 98.08 | 92.73 | 95.33 | 96.97 | 7.27 | 3.03 | 88.32 |
| SRBCT | mRMR-BLO-SVM | 98.13 | 1.87 | 98.80 | 98.80 | 98.80 | 95.83 | 1.20 | 4.17 | 94.63 |
|  | mRMR-BLO-AdaBoost | 98.15 | 1.85 | 98.63 | 98.63 | 98.63 | 97.14 | 1.37 | 2.86 | 95.77 |
|  | mRMR-BLO-ELM | 95.88 | 4.12 | 96.30 | 96.30 | 96.30 | 95.35 | 3.70 | 4.65 | 91.65 |
| Lung Cancer | mRMR-BLO-SVM | 98.15 | 1.85 | 98.81 | 98.81 | 98.81 | 95.83 | 1.19 | 4.17 | 94.64 |
|  | mRMR-BLO-AdaBoost | 98.18 | 1.82 | 98.67 | 98.67 | 98.67 | 97.14 | 1.33 | 2.86 | 95.81 |
|  | mRMR-BLO-ELM | 95.96 | 4.04 | 96.43 | 96.43 | 96.43 | 95.35 | 3.57 | 4.65 | 91.78 |
| Lung(Omic) | mRMR-BLO-SVM | 96.70 | 3.30 | 97.56 | 95.24 | 96.39 | 97.96 | 4.76 | 2.04 | 93.38 |
|  | mRMR-BLO-AdaBoost | 97.80 | 2.20 | 97.92 | 97.92 | 97.92 | 97.67 | 2.08 | 2.33 | 95.59 |
|  | mRMR-BLO-ELM | 95.60 | 4.40 | 94.59 | 94.59 | 94.59 | 96.30 | 5.41 | 3.70 | 90.89 |
| Leukemia(Omic) | mRMR-BLO-SVM | 96.15 | 3.85 | 97.30 | 97.30 | 97.30 | 93.33 | 2.70 | 6.67 | 90.63 |
|  | mRMR-BLO-AdaBoost | 95.92 | 4.08 | 97.78 | 97.78 | 97.78 | 75.00 | 2.22 | 25.00 | 72.78 |
|  | mRMR-BLO-ELM | 94.23 | 5.77 | 97.30 | 94.74 | 96.00 | 92.86 | 5.26 | 7.14 | 85.76 |
| Breast (Omic) | mRMR-BLO-SVM | 92.42 | 7.58 | 94.44 | 91.89 | 93.15 | 93.10 | 8.11 | 6.90 | 84.72 |
|  | mRMR-BLO-AdaBoost | 90.91 | 9.09 | 93.02 | 93.02 | 93.02 | 86.96 | 6.98 | 13.04 | 79.98 |
|  | mRMR-BLO-ELM | 93.94 | 6.06 | 95.74 | 95.74 | 95.74 | 89.47 | 4.26 | 10.53 | 85.22 |

- With an incredible accuracy of 94.44%, the greatest precision for the Colon Tumour dataset was attained by mRMR-BLO-AdaBoost, which outperformed the other models by 3.16% and 1.59%. Reflecting its improved classification accuracy, this model also showed a Precision (PRE) score of 94.23%, Recall (REC) of 98%, F1-score (F1-S) of 96.08%, Specificity (SPE) of 86.36%, and an MCC of 86.76%.
- The mRMR-BLO-ELM had the most outstanding performance in the CNS dataset as compared to mRMR-BLO-AdaBoost (95.60%) by 0.05% and mRMR-BLO-SVM (92.86%). by 2.79% with an accuracy of 97.30%. It noted a recall of 97.30%, F1 score of 97.30%, specificity of 88.89%, and the best MCC of 86.19%.
- Looking at the Ovarian dataset, mRMR-BLO-SVM showed an excellent precision for the classifiers with an unequaled accuracy of 97.56%, exceeding mRMR-BLO-Elm (96.20%) and mRMR-BLO-AdaBoost (95.12%), by 1.41% and 2.56%, respectively. Its classification strength was validated by its comparable metrics, precision, recall, and F-1S of 98.08%, specificity of 96.67%, and MCC of 94.74%.
- For the ALL-AML dataset, the mRMR-BLO-SVM demonstrated an unparalleled accuracy at 97.53%, continuing to exceed mRMR-BLO-Elm stood at 96.20% and mRMR-BLO-AdaBoost 95.12% with a remarkable 1.38% and 2.53% advantage. Its accuracy, recall, and F1-score were noted at 98.08%, with specificity at 96.55% and MCC of 94.63%, underscoring even better classification precision.
- With an accuracy of 95.12%, mRMR-BLO-AdaBoost outperformed mRMR-BLO-SVM and mRMR-BLO-ELM by 1.07% and 8.53%, respectively, for the Breast dataset. It reported the Precision, Recall, F1-score, Specificity, and MCC values of 95.74%, 95.74%, 95.74%, 94.29%, and 90.03%, respectively.
- With an accuracy of 97.89%, mRMR-BLO-SVM excelled among the other participants in the Prostate Tumour dataset. By 0.14% and 2.54%, it also exceeded mRMR-BLO-AdaBoost and mRMR-BLO-ELM. With Precision 98.41%, Recall 98.41%, F1-score 98.41%, Specificity 96.88%, and MCC 95.29% indicating sufficient classification efficiency, the classification metrics fell into the appropriate range.
- Turning to the MLL dataset, mRMR-BLO-AdaBoost outperformed mRMR-BLO-SVM and mRMR-BLO-ELM by 0.22% and 2.38%, respectively, with 97.78% maximum accuracy. Preserving its claim for classification excellence, the computed scores were Precision 98.18%, Recall 98.18%, F1-score 98.18%, Specificity 97.14%, and MCC 95.32%.
- With an accuracy of 98.06%, mRMR-BLO-AdaBoost produced the best results for the Leukemia-3c dataset. It exceeded mRMR-BLO-SVM by 0.48% and mRMR-BLO-ELM by 2.14%, respectively. These findings let the participant attain a Precision of 98.53%, a Recall of 98.53%, an F1-score of 98.53%, a Specificity of 97.14%, and an MCC of 95.67%, so that its classification is not negated.
- The mRMR-BLO-SVM performed 0.03% higher than mRMR-BLO-AdaBoost (96.67%) and 2.53% better than mRMR-BLO-Elm (94.32%) for the Leukemia-4c dataset with best accuracy of 96.70%. Reflecting classification power, it had a Precision of 98.18%, Recall of 96.43%, F1-score of 97.30%, Specificity of 97.14%, and an MCC of 93.10%.
- Similarly, the mRMR-BLO-SVM (98.13%), by 0.02%, and mRMR-BLO-ELM (95.89%), mRMR-BLO-AdaBoost grabbed the lead in the SRBCT dataset with an accuracy of 98.15%. With an MCC of 95.77%, it proved to be strong, showing a Precision of 98.63% as well as Recall, F1-score, and Specificity of 98.63% and 97.14%.
- The best accuracy of 98.18% for the Lung Cancer dataset was with mRMR-BLO-AdaBoost, 0.03% higher than mRMR-BLO-SVM (98.15%) and 2.22% more than mRMR-BLO-Elm (95.96%). Along with an F1-score of 98.67%, Specificity of 97.14%, and MCC of 95.81%, it attained Precision and Recall of 98.67%; the low false positive and false negative rates suggest the great classification capacity for lung cancer diagnosis.
- With the Lung Cancer Omic dataset, the proposed model, mRMR-BLO-AdaBoost, outperforms the two models by showing a maximum accuracy level of 97.80%. The mRMR-BLO-SVM model shows an accuracy level of 97.70%, followed by the mRMR-BLO-ELM model with an accuracy level of 95.60%.
- Using the Leukemia Omic dataset, the proposed mRMR-BLO-SVM shows a maximum accuracy level of 96.15%, followed by the mRMR-BLO-AdaBoost model with an accuracy level of 95.92%, and the mRMR-BLO-ELM with an accuracy level of 94.23%.
- For the Breast Cancer Omic dataset, the mRMR-BLO-ELM is outperforming the other two models, mRMR-BLO-SVM and mRMR-BLO-AdaBoost. The mRMR-BLO-ELM model shows an accuracy of 93.94%, followed by the mRMR-BLO-SVM model of 92.42% accuracy and the mRMR-BLO-AdaBoost model of 90.91% accuracy level.
- The application of SVM, AdaBoost, and ELM classifiers on several datasets using the mRMR-BLO framework is shown in Table 5. With seven out of twelve datasets, mRMR-BLO-AdaBoost clearly attained the greatest accuracy in feature selection and classification—the highest outcome of any strategy investigated. While mRMR-BLO-ELM only excelled in one dataset but has consistent performance in other datasets, mRMR-BLO-SVM fared well in four datasets, indicating high performance in generalizing and resilience. The clear benefit of mRMR-BLO-AdaBoost over the other techniques indicates that the ensemble learning techniques help in raising classification precision and dependability, therefore indicating that the method can properly manage high-dimensional biomedical data. The ROC analysis of the proposed model, in contrast to all datasets, is represented in Figure 2.

### 4.3. Comparative Analysis

- For the Colon Cancer dataset the proposed mRMR-BLO-AdaBoost is achieving the highest accuracy 94.44% which outperforms the [19] by ∼ 5.16%, [20] by ∼ 7.7%, [21] by ∼ 6%, [22] by ∼ 5.33%, and [23] by ∼ 2.21%. However, the other two proposed models, mRMR-BLO-SVM and mRMR-BLO-ELM, show an accuracy of 91.55% and 92.96%, respectively.
- Considering the performance comparison of the proposed model with existing literature in contrast to the CNS dataset, the proposed mRMR-BLO-ELM is the best-performing model with an accuracy of 95.65%. The proposed model outperforms the [24] by ∼ 1%, [25] by ∼ 0.7%, [26] by ∼ 11.32%, [27] by ∼ 7.30%. However, the proposed model mRMR-BLO-ELM lags marginally [28] by ∼ 0.9%. The remaining proposed models, mRMR-BLO-SVM and mRMR-BLO-AdaBoost, show an accuracy of 92.86% and 96.60%.
- Using the Ovarian dataset the proposed mRMR-BLO-SVM shows the highest accuracy of 97.56%, which outperforms citeyaqoob2024optimizing by ∼ 1%, [24] by ∼ 2.7%, [19] by ∼ 0.2%. However, the proposed model mRMR-BLO-ELM underperforms marginally [31] by ∼ 0.07%. The remaining proposed models, mRMR-BLO-AdaBoost and mRMR-BLO-ELM, show an accuracy of 95.12% and 96.20%.
- Considering the performance of the proposed model, mRMR-BLO-SVM shows a maximum accuracy of 97.53% with the ALL-AML dataset. While comparing this with existing literature, the above-mentioned proposed model outperforms [24] by ∼ 1.74%, [32] by ∼ 1%, [29] by ∼ 6.8%, [33] by ∼ 0.4%, and [19] by 1∼ .2%. The other two proposed models, mRMR-BLO-AdaBoost and mRMR-BLO-ELM, show an accuracy of 95.12% and 96.20%, respectively.
- With the Breast Cancer dataset, the maximum performance is shown by the proposed mRMR-BLO-AdaBoost with an accuracy of 95.12%. While comparing this model with some existing literature, the proposed model outperforms [36] with IG and GWO by ∼ 7%, [20] by ∼ 7.6%, [22] ∼ 4.6%, [36] with IG and SVM by ∼ 13.8%, [28] by ∼ 5.56%. The other two proposed models, mRMR-BLO-SVM and mRMR-BLO-ELM, show an accuracy of 94.05% and 86.59%, respectively.
- For the Prostate dataset, the proposed model mRMR-BLO-SVM shows a maximum accuracy of 97.89%. Comparing the proposed model with the existing literature, the proposed model outperforms [33] by ∼ 3.5%, ∼ 1.5%, and ∼ 0.4%, [31] by ∼ 11.5%, and [26] by ∼ 0.9%. The remaining two models, mRMR-BLO with AdaBoost and ELM, show accuracy levels of 97.75% and 95.35%, respectively.
- Using the MLL dataset, the highest accuracy is shown by the proposed model mRMR-BLO-AdaBoost, which has an accuracy of 97.78%. It outperforms [22] by ∼ 16.2%, [25] by ∼ 1.2%, [19] by ∼ 5.3% and [31] by ∼ 29.12%. However, it underperforms [33] by ∼ 1.9%. The other two proposed approaches, including mRMR-BLO with SVM and ELM, show the accuracy level as 97.56% and 95.40%, respectively.
- The proposed mRMR-BLO-AdaBoost shows a maximum accuracy of 98.06% for the Leukemia-3c dataset. While comparing the performance of the above-mentioned approach to the existing literature, it outperforms [31] by ∼ 4.4%, [19] by ∼ 4.14%, [34] by ∼ 3.6%, and [25] marginally by ∼ 0.09%. The other two approaches, including mRMR-BLO-SVM and mRMR-BLO-ELM, show an accuracy level of 97.59% and 95.92%, respectively.
- With the Leukemia-4c dataset, the proposed mRMR-BLO-SVM shows a maximum accuracy level of 96.70%. When it is compared to the performance of the existing literature, the proposed model outperforms the [19] by ∼ 6%, [34] by ∼ 6%, and [25] by ∼ 2.46%. While comparing the performance with [35], the proposed model lags by ∼ 0.4%. The other two proposed models, including mRMR-BLO-AdaBoost and mRMR-BLO-ELM, show an accuracy level of 96.67% and 94.32%, respectively.
- For the SRBCT dataset, the proposed mRMR-BLO-AdaBoost shows a maximum accuracy of 98.15%. It outperforms the existing literature including [20] by ∼ 1.7%, [34] by ∼ 2.4%, [31] by ∼ 3%, [27] by ∼ 1%. In addition, the other two proposed approaches, including mRMR-BLO-SVM and mRMR-BLO-ELM, show an accuracy of 98.13% and 95.88%, respectively.
- With the Lung Cancer dataset, the proposed mRMR-BLO-AdaBoost shows a maximum accuracy of 98.18%. It outperforms the [37] by ∼ 18.5%, [24] by ∼ 4.2%, [20] by ∼ 4%, [30] by ∼ 2.1%. While comparing to [26], the proposed model lags marginally by ∼ 0.4%. The other proposed approaches, including mRMR-BLO-SVM and mRMR-BLO-ELM, show an accuracy level of 98.15% and 95.96%, respectively.
- Table 6 shows the comparative study analysis done for the proposed models with some existing literature. The accuracy comparison of the proposed model with existing literature, in contrast to different datasets, is represented in Figure 3.

**Table 6:**
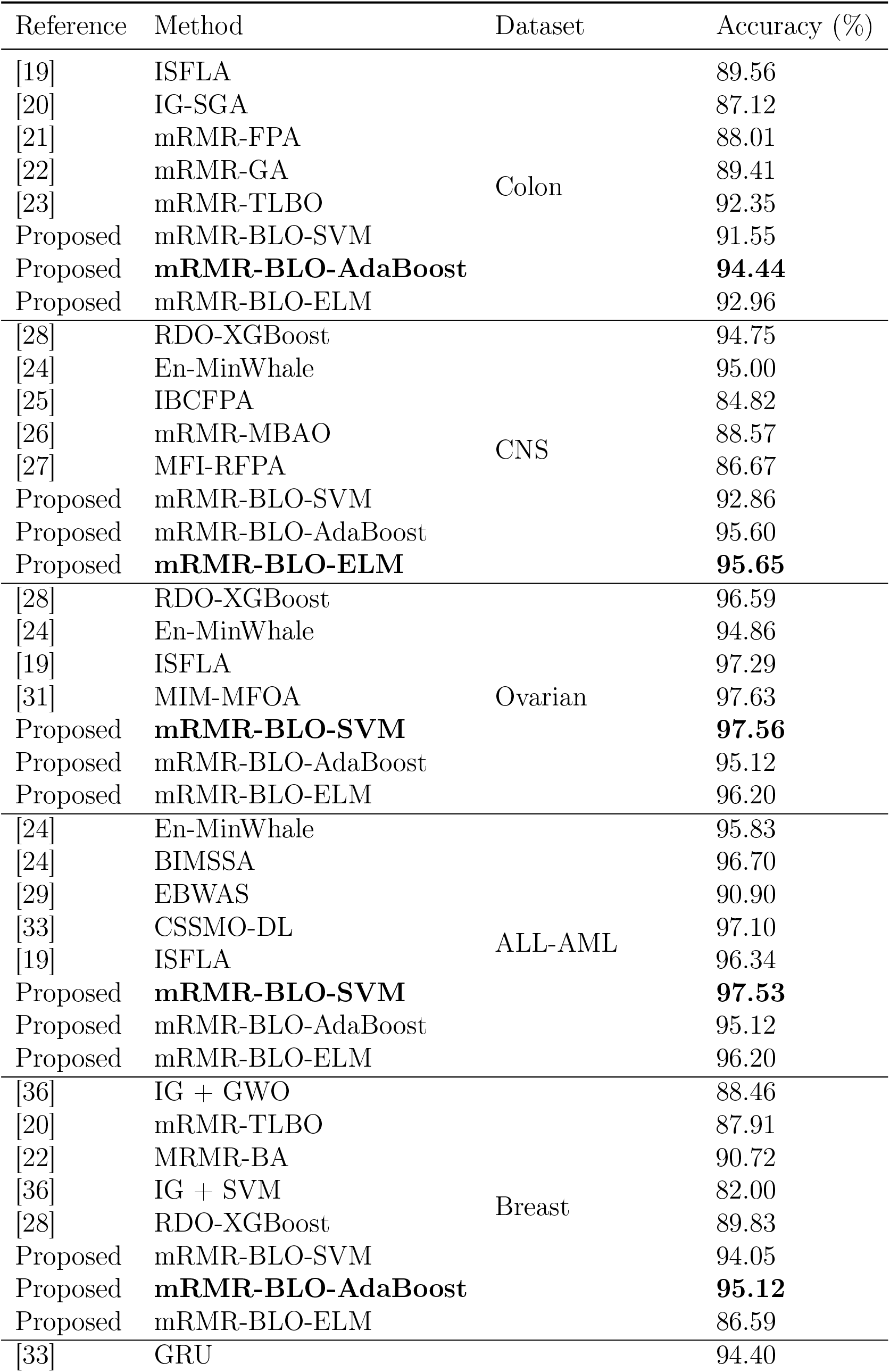
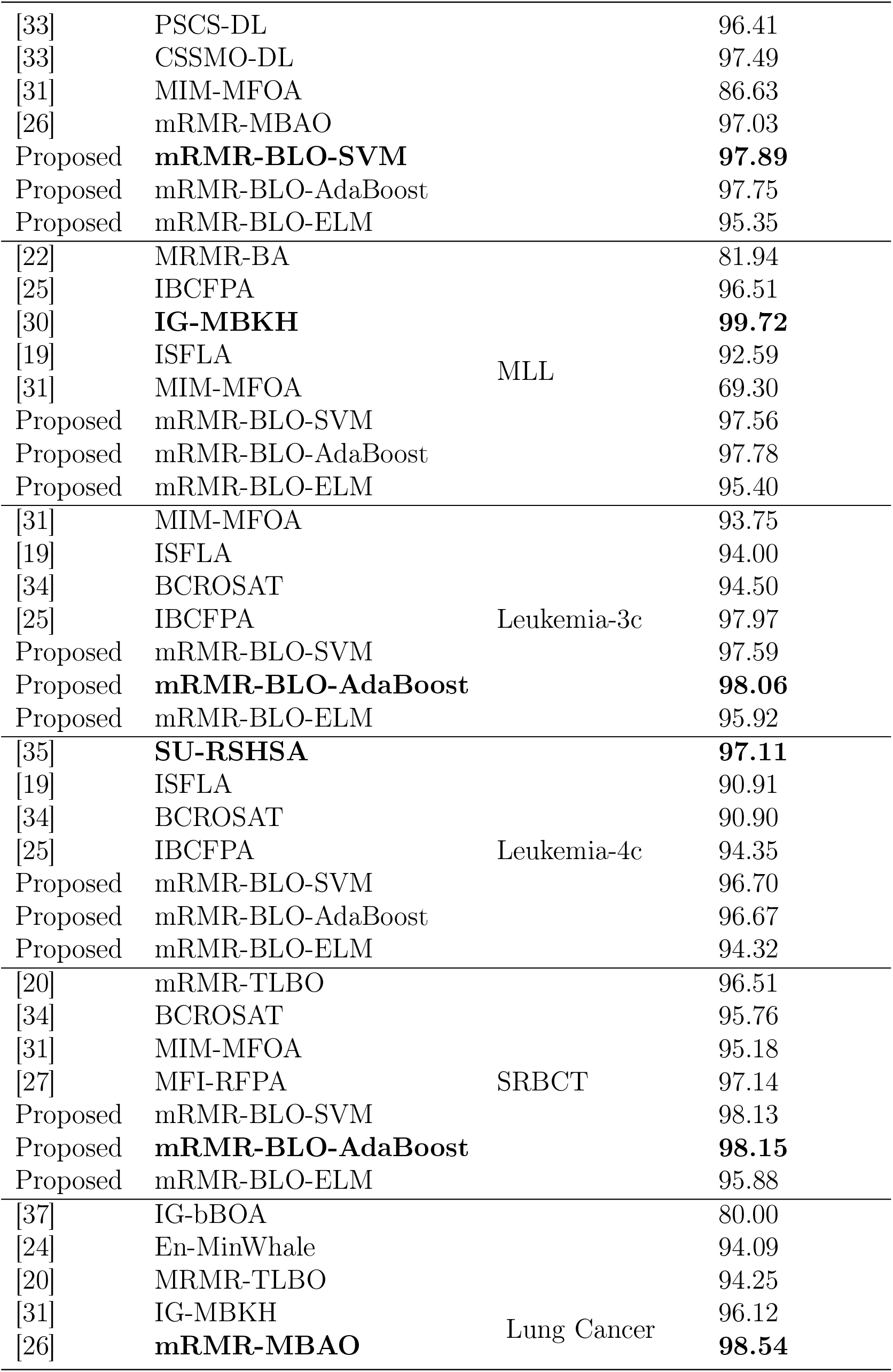
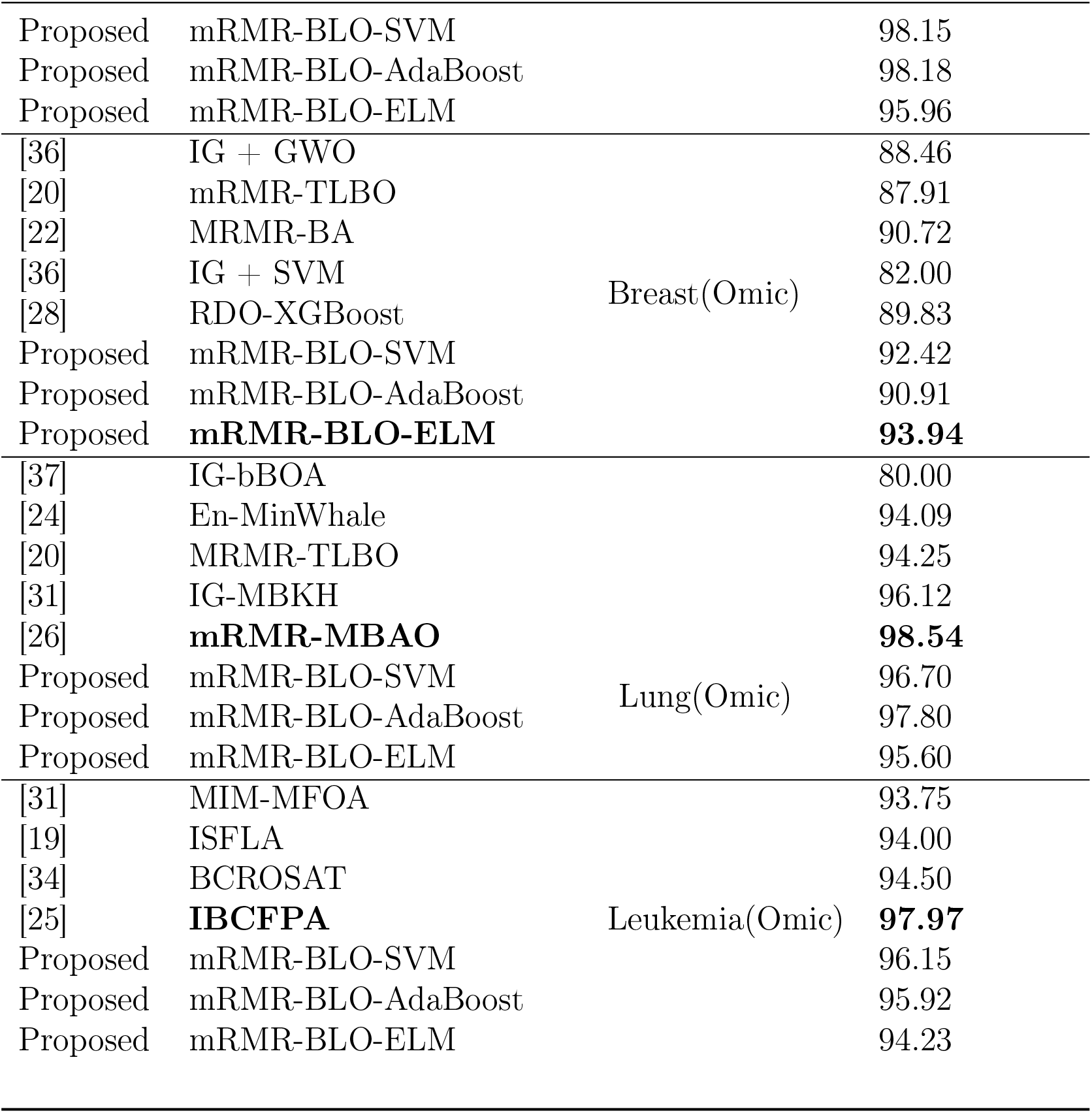
Comparison of classification accuracy across different methods and datasets.

**Figure 3:**
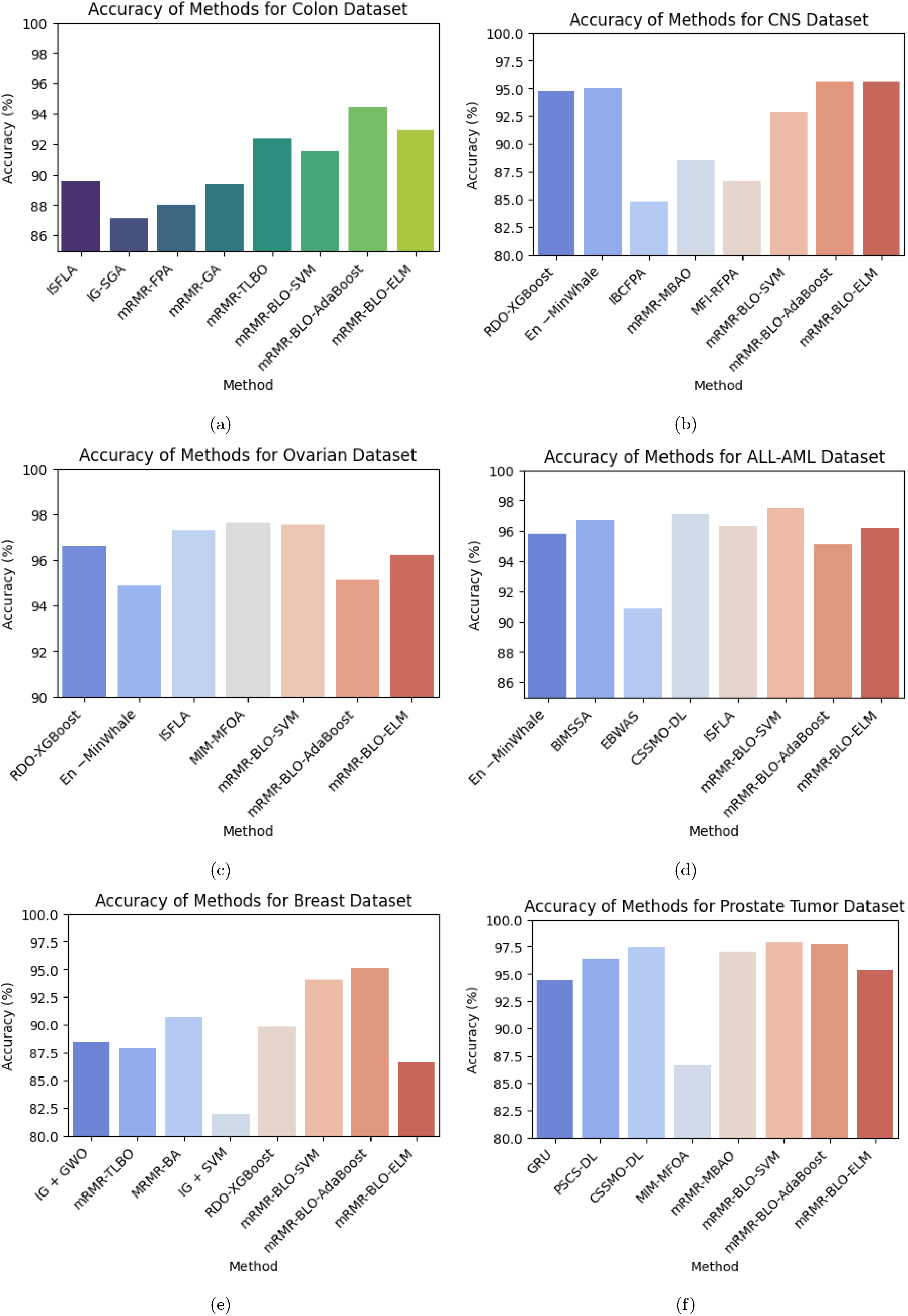
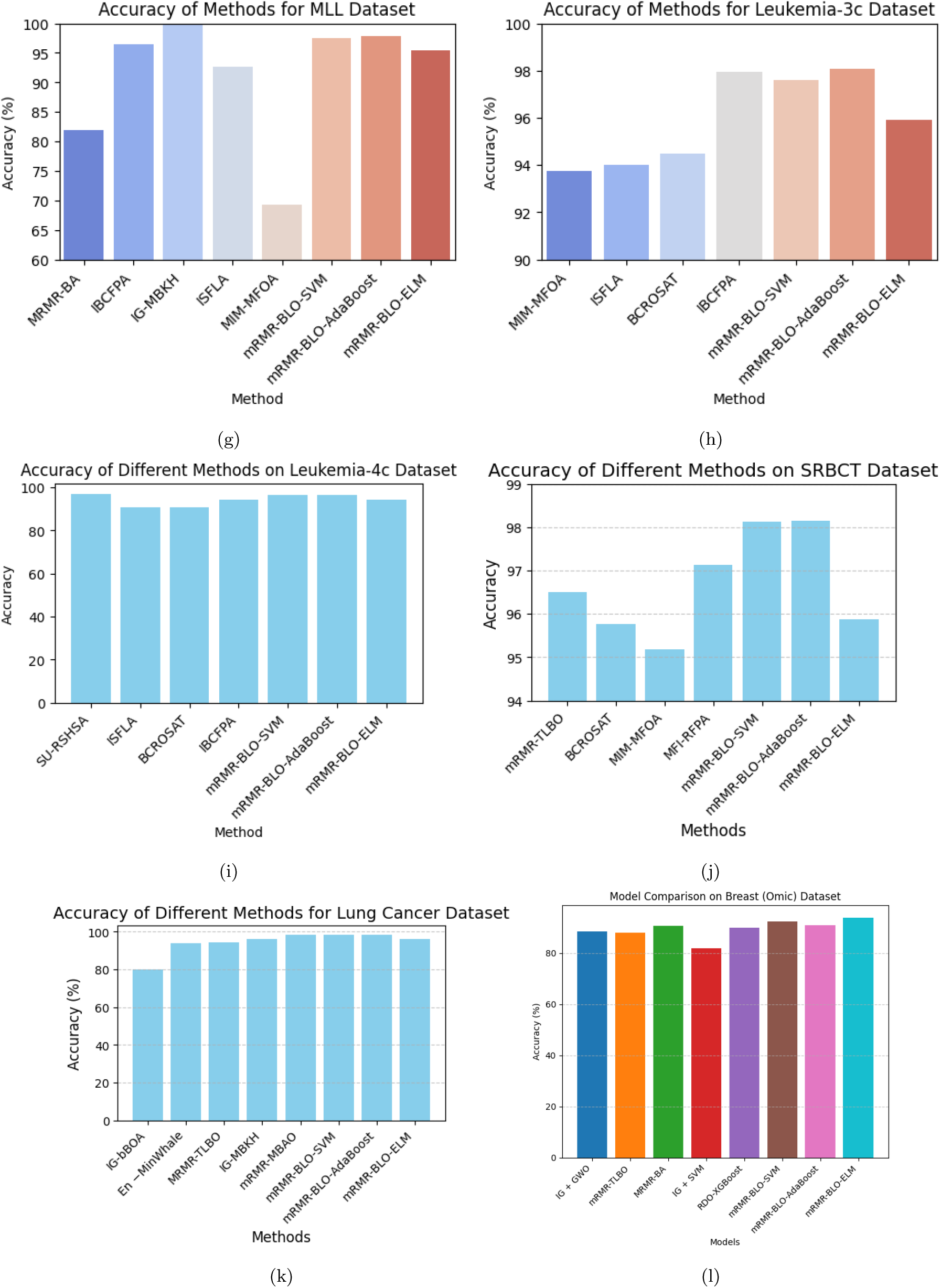
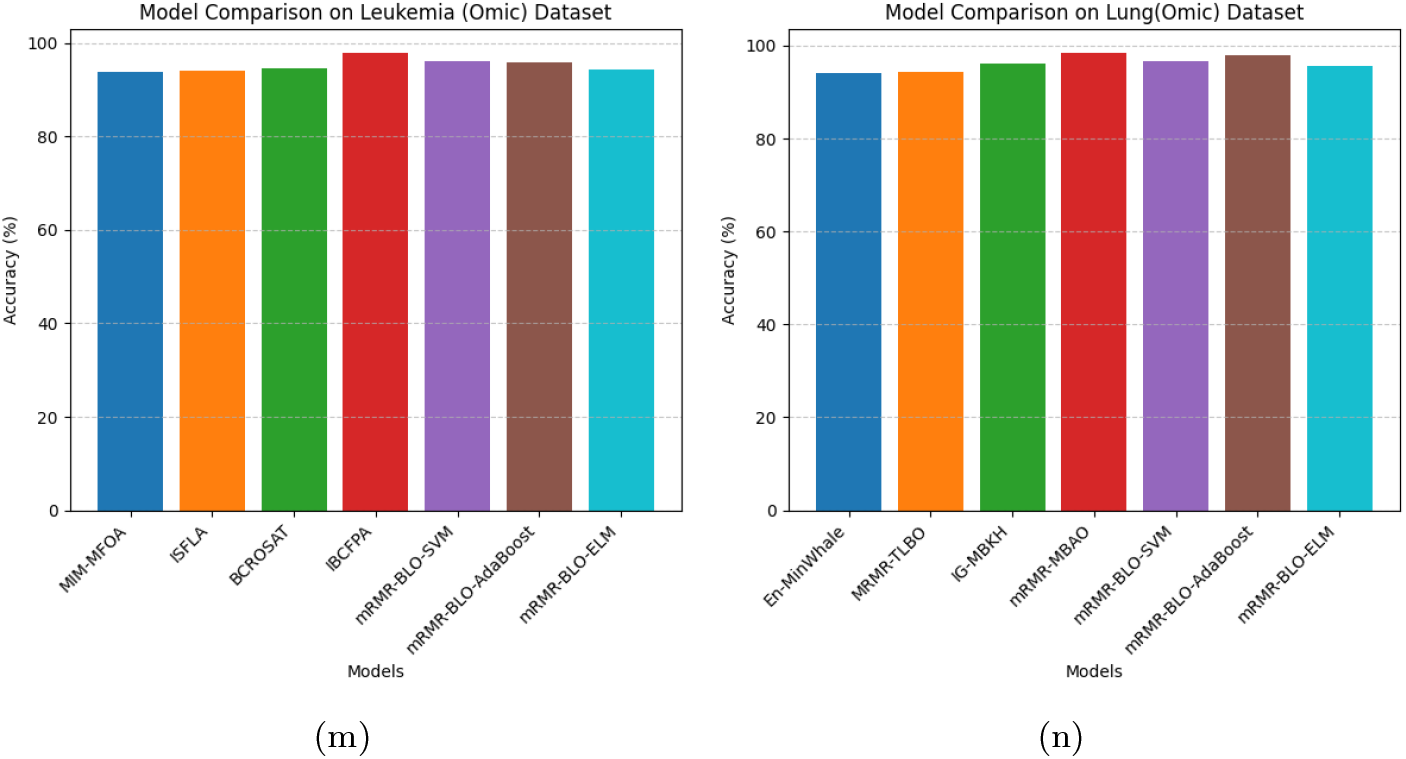
Accuracy comparison of proposed models with existing literature incontrast to (a) Colon Tumor CNS (c) Ovarian (d) ALL-AML (e) Breast Cancer (f) Prostate Tumor (g) MLL (h) Leukemia-3c (i) Leukemia-4c (j) SRBCT (K) Lung Cancer (l) Breast(Omic) (m) Luekemia(Omic) (n) Lung(Omic) Dataset

### 4.4. Comparison of mRMR-BLO with other algorithms using the Wilcoxon paired signed rank test

We investigated whether mRMR-BLO performs at the same level as other techniques using the nonparametric Wilcoxon Paired Signed Ranks test. Although the alternative hypothesis (*H*_1_) shows that our method shows a statistically significant difference over other FS algorithms, the null hypothesis of this test (*H*_0_) argues that there is no appreciable improvement over other gene selection algorithms.

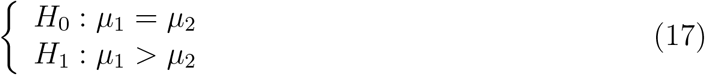

Equation 17 shows the null hypothesis (*H*_0_) and the alternative hypothesis (*H*_1_), where, respectively, the average accuracy from our method and other algorithms on all datasets. In this study, we have utilized Paired Wilcoxon’s signed rank test among mRMR-BLO with some additional algorithms, including mRMR-MBAO [26], IG-MBKH[30], MIM-MFOA [31], gene, and IBCFPA [25].

Table 7 shows the accuracy with Wilcoxon’s signed rank test of mRMR-BLO with these methods for the MLL dataset. Table 7 shows that among mRMR-BLO and other methods, the p-values derived by the paired Wilcoxon signed rank test all lie below the five percent standard significance level. As such, we embrace the alternative hypothesis (*H*_1_) while rejecting the null hypothesis (*H*_0_). Our method (mRMR-BLO) thus beats many other methods. MRMR-BLO has been successful and efficient. Consequently, we find that mRMR-BLO offers superior solutions to other algorithms. Our method differs significantly from existing comparison methods such as mRMR-MBAO, IG-MBKH, MIM-MFOA, and IBCFPA.

**Table 7:** Paired Wilcoxon’s signed-rank test results comparing mRMR-BLO with other methods.

| Paired Wilcoxon's Test | mRMR-BLO vs |  |  |  |
| --- | --- | --- | --- | --- |
|  | mRMR-MBAO | IBCFPA | IG-MBKH | MIM-MFOA |
| *p*-Value |  |  |  |  |
| Accuracy | 0.00861 | 0.03341 | 0.01154 | 0.01725 |

## 5. Conclusions

In machine learning, dimensionality reduction is absolutely important because it reduces model complexity, increases accuracy, increases computing efficiency, and helps to prevent overfitting. This paper presents the binary variant of the current LO algorithm as a wrapper-based FS method coupled with mRMR to exploit these benefits. Based on its simplicity and efficiency, the suggested approach is evaluated on eleven cancer datasets using the KNN classifier. Among various well-known FS methods, including mRMR-MBAO, IBPSO, IG-MBKH, and MIM-MFOA, mRMR-BLO is evaluated. With regard to certain evaluation criteria, model stability is investigated using the Paired Wilcoxon signed rank test. With the best classification accuracy of 98.15% attained using mRMR-BLO-AdaBoost on the SRBCT dataset, the results show the excellence of the suggested strategy. Furthermore, the model does really well on other settings. We evaluate its performance using three classifiers: ELM, AdaBoost, and SVM. MRMR-BLO might eventually find use in practical fields, including picture encryption and human activity detection. Hybridizing it with another metaheuristic, such as Simulated Annealing (SA), could provide even more improvements.

## Declarations

### Ethics approval and consent to participate

Not Applicable.

### Consent to Publication

Not applicable. The manuscript does not include any individual person’s data in any form requiring consent.

## Data Availability

Data is available on 11 publicly available microarray gene expression datasets and the Omic cancer dataset for different diseases, https://data.mendeley.com/datasets/fhx5zgx2zj/ 1 and https://sbcb.inf.ufrgs.br/cumida#datasets respectively.

